# Association of the inflammation-related proteome with dementia development at older age: results from a large, prospective, population-based cohort study

**DOI:** 10.1101/2021.06.15.21258913

**Authors:** Kira Trares, Megha Bhardwaj, Laura Perna, Hannah Stocker, Agnese Petrera, Stefanie M. Hauck, Konrad Beyreuther, Hermann Brenner, Ben Schöttker

**Affiliations:** Network Aging Research, Heidelberg University, Bergheimer Straße 20, 69115 Heidelberg, Germany; Division of Clinical Epidemiology and Aging Research, German Cancer Research Center, Im Neuenheimer Feld 581, 69120 Heidelberg, Germany; Medical Faculty, Heidelberg University, Im Neuenheimer Feld 672, 69120 Heidelberg; Department of Translational Research in Psychiatry, Max Planck Institute of Psychiatry, Kraepelinstraße 2-10, 80804 Munich, Germany; Division of Mental Health of Older Adults, Department of Psychiatry and Psychotherapy, University Hospital, LMU Munich, 80336 Munich, Germany; Research Unit Protein Science and Metabolomics and Proteomics Core Facility, Helmholtz Zentrum Munich - German Research Center for Environmental Health, Heidemannstraße 1, 80939 Munich

## Abstract

**Importance:** Chronic inflammation is increasingly recognized as a central feature of several forms of dementia.

**Objective:** To determine which biomarkers of the inflammation-related proteome are associated with all-cause dementia, Alzheimer’s disease (AD), or vascular dementia (VD).

**Design:** Analyses were performed in a case-cohort study design based on an ongoing German population-based cohort study.

**Setting:** Serum samples of study participants were collected at baseline (2000-20002), and participants were followed up for 17 years. Information about a dementia diagnosis was collected during follow-up via collection of medical records from general practitioners.

**Participants:** Ascertainment of potential dementia development during follow-ups was conducted for 6,284 study participants aged 50-75 years at baseline. Biomarker measurements were performed in a randomly collected sample of 1,435 participants and all incident dementia cases of the rest of the cohort (n=393).

**Main Outcomes and Measures:** All-cause dementia, AD and VD were the primary outcomes of this analysis.

**Results:** Biomarkers were analyzed in 504 all-cause dementia cases (mean age, 67.0 [SD, 5.1] years; 262 female [52.0%], and 242 male [48.0%]) and 1,278 controls (mean age, 61.9 [standard deviation (SD): 6.5] years; 703 female [55.0%], and 575 male [45.0%]). Among the dementia cases, 163 participants developed AD and 195 VD. After correction for multiple testing, 58 biomarkers were statistically significantly associated with all-cause dementia, 22 with AD, and 33 with VD incidence. All analyses were adjusted for potential confounders. Besides single biomarker associations, we identified four biomarker clusters based on the strongest and independently associated biomarkers CX3CL1, EN-RAGE, LAP TGF-beta-1 and VEGF-A. CX3CL1 (Odds ratio [95%-confidence interval] per 1 standard deviation increase: 1.41 [1.24-1.60]) and EN-RAGE (1.41 [1.25-1.60]) were associated with all-cause dementia incidence, EN-RAGE (1.51 [1.25-1.83]) and LAP TGF-beta-1 (1.46 [1.21-1.76]) with AD incidence, and VEGF-A (1.43 [1.20-1.70]) with VD incidence. All named associations were stronger among *APOE* ε4 negative subjects.

**Conclusion and Relevance:** This study shows for the first time that the majority of inflammation-related proteins measured in serum samples (58 of 72 tested (80.6%)) are associated with all-cause dementia incidence. Future studies should not only concentrate on single biomarkers but also the complex relationships in biomarker clusters.

**Key Points:** *Question:* Which biomarkers of the inflammatory proteome are risk factors for dementia?

*Findings:* After correction for multiple testing, in this large prospective cohort study (n=1,782), 58 of 72 tested (80.6%) inflammation-related proteins were associated with all-cause dementia. Furthermore, 22 and 33 were significantly associated with Alzheimer’s disease and vascular dementia. Due to high inter-correlation, only four biomarkers (CX3CL1, EN-RAGE, LAP TGF-beta-1, VEGF-A) were independently associated with dementia outcomes.

*Meaning:* The underlying pathophysiology of dementia development might involve complex inflammatory protein clusters, and the identified biomarkers might be promising new drug targets, early diagnostic markers, or parts of prediction models.

## 1. Introduction

Dementia is a major challenge for global public health and social care systems. As the number of dementia cases increases with rising life expectancy, it has been estimated that almost 75 million people worldwide will live with dementia in 2030^1^. Therefore, research on how to prevent or delay the onset of dementia is one of the major challenges globally^2^.

Inflammation likely plays a key role in the development and progression of dementia^3^. In the brain, inflammatory processes are a defence mechanism against infection, toxins, or injury. However, persistent inflammation might disrupt the equilibrium of pro- and anti-inflammatory signalling. In neuroinflammation, microglia and astrocytes are activated and release various pro-inflammatory products causing chronic inflammation and neurodegeneration^4, 5^. Several studies have found increased levels of pro-inflammatory cytokines and proteins like interleukin-6 (IL-6), interleukin-1β (IL-1β), C-reactive protein (CRP), or α1-antichymotrypsin (α1-AT) to be associated with the onset of all-cause dementia^6, 7^. Further studies also revealed that CRP, IL-1ß, IL-2, IL-4, IL-6, IL8, IL-10, IL-12, IL-18, monocyte protein-1 (MCP-1), MCP-3, interferon-γ-inducible protein 10 (IP-10), and tumour necrosis factor α (TNF-α) are associated with the incidence of Alzheimer’s disease (AD), the most common form of dementia, accounting for 60-80% of all cases^8, 9^. However, longitudinal studies on the association between biomarkers of the inflammation-related proteome and dementia are scarce^10^.

As biomarkers can be measured at an early stage of dementia, they have the potential to be used for an early diagnosis^11, 12^. Furthermore, identified new biomarkers could deepen our understanding of the pathogenetic processes leading to dementia and might represent novel drug targets^13, 14^. The current challenge of biomarker research in dementia in general and distinct forms is to find reliable diagnostic and predictive biomarkers easily accessible in fluids like blood^15, 16^.

This study aims for the first time to identify blood-based biomarkers from a set of 92 inflammatory biomarkers as risk factors for all-cause dementia, Alzheimer’s disease (AD), or vascular dementia (VD) incidence in a large, prospective cohort study with a 17-year follow-up.

## 2. Methods

### 2.1. **Study population**

We conducted a case-cohort study based on the ESTHER study (Epidemiologische Studie zu Chancen der Verhütung, Früherkennung und optimierten Therapie chronischer Erkrankungen in der älteren Bevölkerung [German]). In this prospective cohort study implemented in Saarland, Germany, 9,940 women and men aged 50 to 75 years at baseline were recruited during a general health checkup by their general practitioners between 2000 and 2002. Besides an age of 50-75 years, the inclusion criteria for the ESTHER study were physical and mental ability to participate in the study as well as knowledge of the German language. Participants were followed up concerning the incidence of major diseases and mortality 2, 5, 8, 11, 14, and 17 years after baseline. For details, see Löw et al. 2004^17^. The sociodemographic baseline characteristics and common prevalent chronic diseases were similarly distributed in the respective age categories as in the German National Health Survey, a representative sample of the German population^17^. The study was approved by the ethics committees of Heidelberg University and the state medical board of Saarland, Germany.

### 2.2. Dementia ascertainment

Information about a dementia diagnosis was collected during the 14- and 17-year-follow-ups of the ESTHER study. The median follow-up time was 16.3 years (interquartile-range: 13.5-17.0 years), and the maximum was 19.4 years due to the 2-year period of baseline recruitment. In brief, the collection of dementia diagnoses included sending standardized questionnaires to the study participants’ general practitioners (GPs). Participants who had dropped out during previous follow-ups due to ill health or had died were included in the dementia ascertainment through the GPs as well. If the GPs were aware of a dementia diagnosis for their patients, they were asked to provide all available medical records documenting a dementia diagnosis. The latter included records from neurologists, psychiatrists, memory clinics, or other specialized providers. If the GP provided a mixed dementia diagnosis, available medical records were screened for an underlying AD or VD background and considered as AD, VD, or both. The current guidelines in Germany for AD diagnosis follow the National Institute on Aging and the Alzheimer’s Association^18^ or the International Working Group (IWG)-2 criteria^19, 20^.

### 2.3. Biomarker and covariate assessment

The serum of baseline blood samples was analyzed with the Olink Target 96 Inflammation panel, developed by Olink Proteomics (Uppsala, Sweden). The details for the biomarker and covariate assessment are shown in **Supplemental Text 1**.

### 2.4. In- and exclusion criteria

The selection of study participants from the ESTHER cohort for this case-cohort analysis is shown in Supplemental Figure 1. ESTHER participants were eligible for selection as cases or random controls. Participants were excluded if dementia incidence status could not be ascertained by GP questionnaires (n=3,583) or blood samples were not available (n=73). Thus, information from 6,284 participants was available for analyses. Olink inflammation panel measurements were performed in a case-cohort design among 1,435 randomly selected study participants and all incident dementia cases of the rest of the cohort (n=393). After excluding participants with quality control warning, 389 incident dementia cases and 1,393 randomly selected participants were available. As the random controls included 115 incident dementia cases, the study population comprised 504 participants with incident dementia and 1,278 randomly selected controls.

### 2.5. Statistical analyses

First, to describe factors associated with dementia risk, categorized baseline characteristics of all-cause dementia cases and controls were compared using the χ^2^-test. Second, odds ratios (ORs) were estimated with a multivariate logistic regression model, including all baseline characteristics.

In a univariate, descriptive analysis, the median and interquartile range (IQR) of all inflammation-related protein levels of all-cause dementia, AD, and VD cases were separately compared with those of controls, using the Wilcoxon Rank Sum test. Additionally, in a multivariate approach, the ORs per one standard deviation (SD) increase of each inflammation-related protein were assessed separately with each outcome (all-cause dementia, AD, and VD incidence) in logistic regression models adjusted for potential confounders. In models for AD incidence, study participants with other (e.g. VD) or unknown dementia forms were excluded. The same was applied for the outcome VD incidence by excluding AD and other non-VD cases. The models were adjusted for age, sex, education, physical activity, BMI, CVD, diabetes, depression, and *APOE* genotype. All variables were used as categorical variables, as described in Table 1, except age, which was modelled continuously. The covariates were selected because they were statistically significantly associated with all-cause dementia, AD, or VD in a previous analysis with the ESTHER study participants^21^. Statistical test results were corrected for multiple testing by the Benjamini and Hochberg method for all tests carried out for one outcome^22^. A false discovery rate (FDR) < 0.05 was applied as the threshold for statistical significance.

**Table 1.**
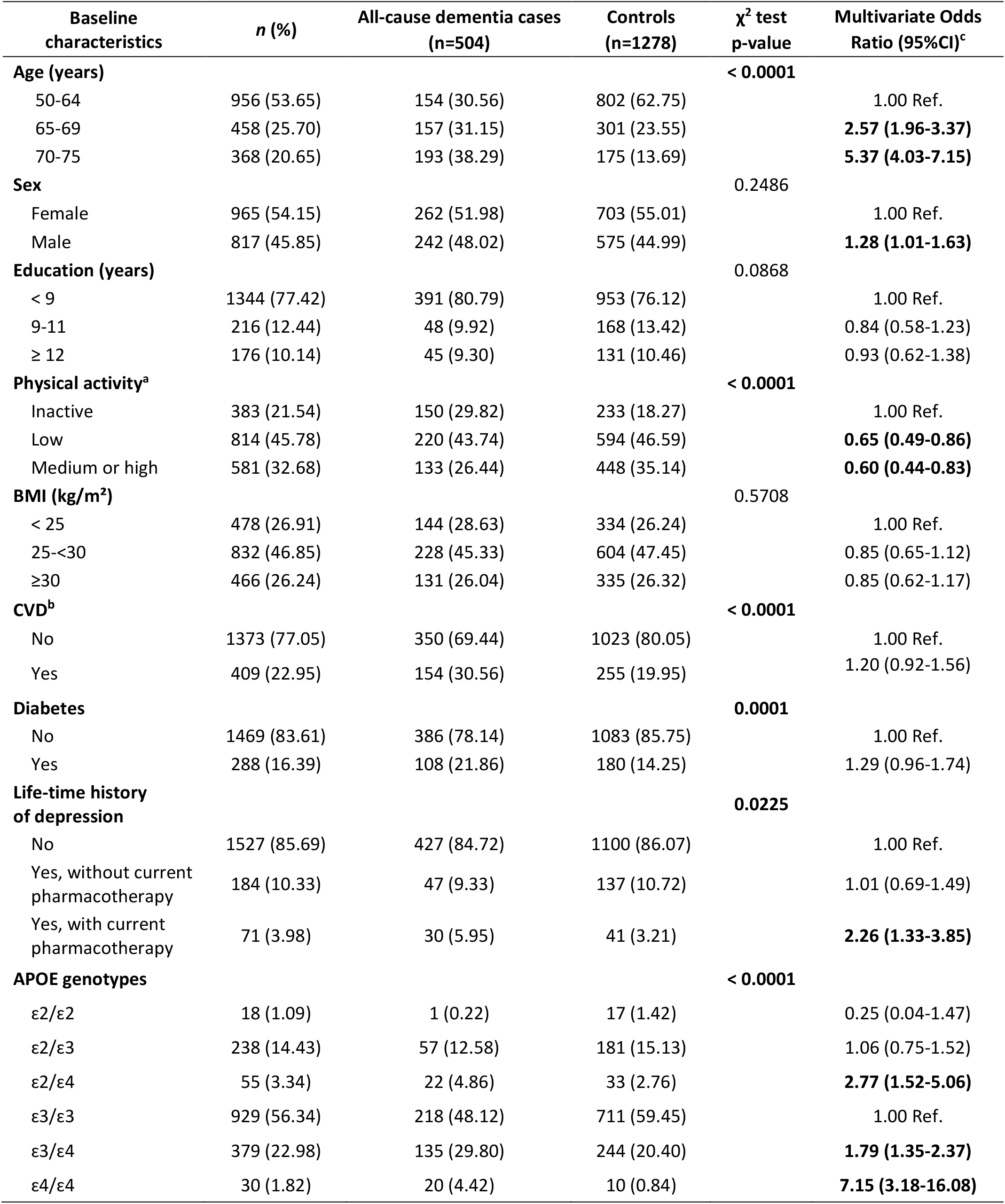
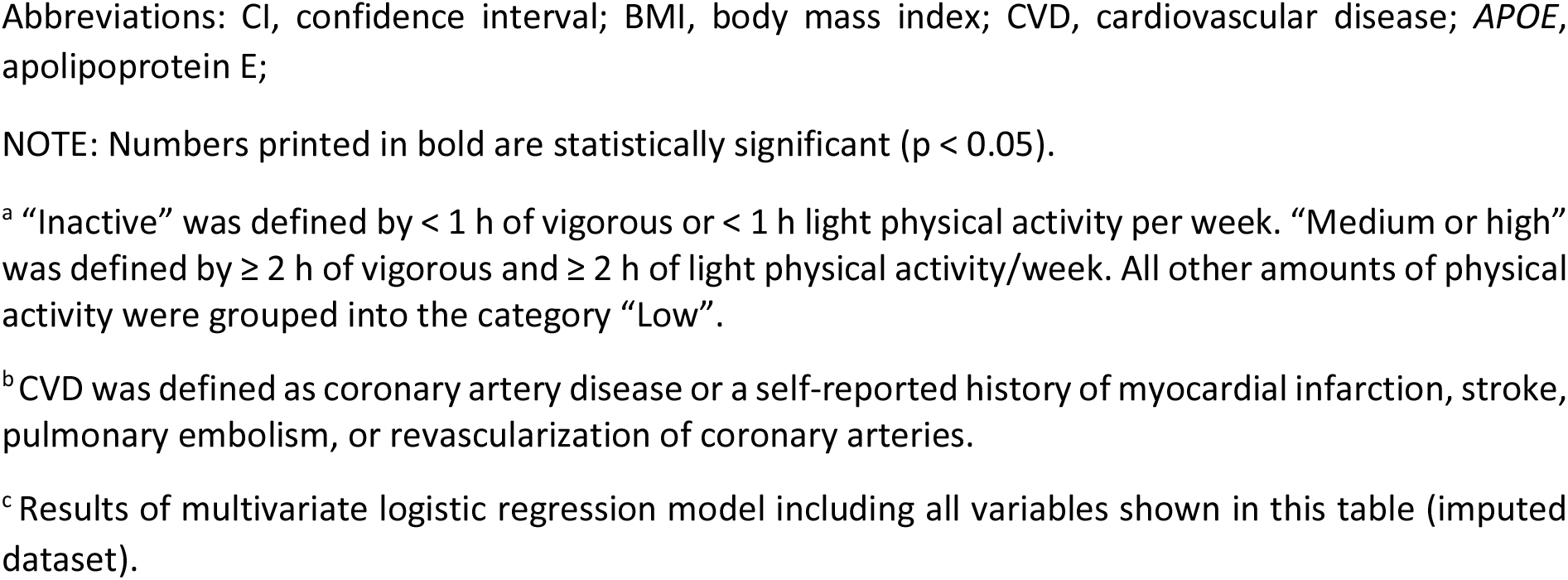
Baseline characteristics of included study participants (n=1782) pharmacotherapy

| Baseline characteristics | n (%) | All-cause dementia cases (n=504) | Controls (n=1278) | $\chi^2$ test p-value | Multivariate Odds Ratio (95%CI)<sup>c</sup> |
| --- | --- | --- | --- | --- | --- |
| **Age (years)** |  |  |  | **< 0.0001** |  |
| 50-64 | 956 (53.65) | 154 (30.56) | 802 (62.75) |  | 1.00 Ref. |
| 65-69 | 458 (25.70) | 157 (31.15) | 301 (23.55) |  | **2.57 (1.96-3.37)** |
| 70-75 | 368 (20.65) | 193 (38.29) | 175 (13.69) |  | **5.37 (4.03-7.15)** |
| **Sex** |  |  |  | 0.2486 |  |
| Female | 965 (54.15) | 262 (51.98) | 703 (55.01) |  | 1.00 Ref. |
| Male | 817 (45.85) | 242 (48.02) | 575 (44.99) |  | **1.28 (1.01-1.63)** |
| **Education (years)** |  |  |  | 0.0868 |  |
| < 9 | 1344 (77.42) | 391 (80.79) | 953 (76.12) |  | 1.00 Ref. |
| 9-11 | 216 (12.44) | 48 (9.92) | 168 (13.42) |  | 0.84 (0.58-1.23) |
| ≥ 12 | 176 (10.14) | 45 (9.30) | 131 (10.46) |  | 0.93 (0.62-1.38) |
| **Physical activity<sup>a</sup>** |  |  |  | **< 0.0001** |  |
| Inactive | 383 (21.54) | 150 (29.82) | 233 (18.27) |  | 1.00 Ref. |
| Low | 814 (45.78) | 220 (43.74) | 594 (46.59) |  | **0.65 (0.49-0.86)** |
| Medium or high | 581 (32.68) | 133 (26.44) | 448 (35.14) |  | **0.60 (0.44-0.83)** |
| **BMI (kg/m<sup>2</sup>)** |  |  |  | 0.5708 |  |
| < 25 | 478 (26.91) | 144 (28.63) | 334 (26.24) |  | 1.00 Ref. |
| 25-<30 | 832 (46.85) | 228 (45.33) | 604 (47.45) |  | 0.85 (0.65-1.12) |
| ≥30 | 466 (26.24) | 131 (26.04) | 335 (26.32) |  | 0.85 (0.62-1.17) |
| **CVD<sup>b</sup>** |  |  |  | **< 0.0001** |  |
| No | 1373 (77.05) | 350 (69.44) | 1023 (80.05) |  | 1.00 Ref. |
| Yes | 409 (22.95) | 154 (30.56) | 255 (19.95) |  | 1.20 (0.92-1.56) |
| **Diabetes** |  |  |  | **0.0001** |  |
| No | 1469 (83.61) | 386 (78.14) | 1083 (85.75) |  | 1.00 Ref. |
| Yes | 288 (16.39) | 108 (21.86) | 180 (14.25) |  | 1.29 (0.96-1.74) |
| **Life-time history of depression** |  |  |  | **0.0225** |  |
| No | 1527 (85.69) | 427 (84.72) | 1100 (86.07) |  | 1.00 Ref. |
| Yes, without current pharmacotherapy | 184 (10.33) | 47 (9.33) | 137 (10.72) |  | 1.01 (0.69-1.49) |
| Yes, with current pharmacotherapy | 71 (3.98) | 30 (5.95) | 41 (3.21) |  | **2.26 (1.33-3.85)** |
| **APOE genotypes** |  |  |  | **< 0.0001** |  |
| ε2/ε2 | 18 (1.09) | 1 (0.22) | 17 (1.42) |  | 0.25 (0.04-1.47) |
| ε2/ε3 | 238 (14.43) | 57 (12.58) | 181 (15.13) |  | 1.06 (0.75-1.52) |
| ε2/ε4 | 55 (3.34) | 22 (4.86) | 33 (2.76) |  | **2.77 (1.52-5.06)** |
| ε3/ε3 | 929 (56.34) | 218 (48.12) | 711 (59.45) |  | 1.00 Ref. |
| ε3/ε4 | 379 (22.98) | 135 (29.80) | 244 (20.40) |  | **1.79 (1.35-2.37)** |
| ε4/ε4 | 30 (1.82) | 20 (4.42) | 10 (0.84) |  | **7.15 (3.18-16.08)** |

Abbreviations: CI, confidence interval; BMI, body mass index; CVD, cardiovascular disease; *APOE*, apolipoprotein E;
NOTE: Numbers printed in bold are statistically significant ( $p < 0.05$ ).
<sup>a</sup> “Inactive” was defined by $< 1$ h of vigorous or $< 1$ h light physical activity per week. “Medium or high” was defined by $\geq 2$ h of vigorous and $\geq 2$ h of light physical activity/week. All other amounts of physical activity were grouped into the category “Low”.
<sup>b</sup> CVD was defined as coronary artery disease or a self-reported history of myocardial infarction, stroke, pulmonary embolism, or revascularization of coronary arteries.
<sup>c</sup> Results of multivariate logistic regression model including all variables shown in this table (imputed dataset).

We further aimed to identify those inflammation-related proteins whose association with a dementia outcome was independent of other inflammatory biomarkers. Therefore, all biomarkers, which were significantly associated with a dementia endpoint after FDR correction, were tested for the independence of the association by forward elimination. In detail, only biomarkers having the strongest, independent, positive association with the outcome entered the regression model with the threshold for statistical significance of p<0.05. The identified independent biomarkers were used for the naming of biomarker clusters. All other biomarkers of the Olink inflammation panel, which were highly correlated (Spearman’s correlation coefficient r > 0.5)^23^ with an independent biomarker, were put in its cluster. One biomarker might be in more than one cluster. We favoured this statistical approach over a principal component analysis (PCA) because it has a higher transparency, is easier to reproduce by others, its results are easier to interpret, and the associations of the biomarkers with the dementia outcomes are being acknowledged in the decision about the number of clusters.

The associations of the independent biomarkers with dementia endpoints were further analyzed in subgroup analyses based on age, sex, obesity, diabetes, history of CVD, and *APOE* ε4 polymorphism. Apart from this, interaction terms were tested. In addition, the dose-response relationships between the independent biomarkers and dementia endpoints were assessed with restricted cubic spline curves^24^. Information on sensitivity analyses and the imputation of missing covariate values are given in **Supplemental Text 2**.

## 3. Results

Table 1 shows the baseline characteristics of 504 cases with incident dementia from any cause and 1,278 controls. The χ^2^ test revealed significant differences between cases and controls in terms of age, physical activity, cardiovascular disease (CVD), diabetes, lifetime history of depression, and *APOE* genotype. In the multivariate logistic regression analysis, CVD and diabetes lost statistical significance, but a trend towards an increased dementia risk could still be seen in the OR point estimates. Age, sex, lifetime history of depression with current pharmacotherapy and having at least one ε4 allele of the *APOE* gene remained significantly positively associated with all-cause dementia incidence. In contrast, physical activity remained significantly inversely associated.

Among the included 504 all-cause dementia cases, 163 and 195 participants developed AD and VD, respectively. The medians of all inflammation-related protein levels of all-cause dementia, AD, and VD cases were separately compared with those of controls (Supplemental Tables 2-4). In this univariate analysis, n = 60, n = 51 and n = 52 biomarker levels of the Olink inflammation panel were significantly increased in all-cause dementia, AD, and VD cases, respectively (FDR < 0.05).

Tables 2-4 show the multivariate logistic regression model results for those 58, 22, and 33 biomarkers, significantly associated with all-cause dementia, AD, and VD incidence, respectively, after FDR correction. Supplemental Tables 5-7 show the non-significant ones. The associations’ strengths were comparable and ranged for the various biomarker-outcome associations from OR point estimates of 1.12 to 1.51 per 1 SD increase. The forward selection revealed that only two (CX3CL1 and EN-RAGE), two (EN-RAGE and LAP TGF-beta-1), and one (VEGF-A) inflammation-related proteins were independently, positively associated with all-cause dementia, AD, and VD, respectively. The reason for the low number of independent inflammation biomarkers was mainly due to high inter-correlation. Overall, 18, 26, 16, and 28 biomarkers of the Olink inflammation panel had a Spearman’s r > 0.5 with CX3CL1, EN-RAGE, LAP TGF-beta 1, and VEGF-A, respectively (Supplemental Tables 8-11).

**Table 2.** Associations of significantly associated Olink Biomarker levels with **all-cause dementia** incidence.

| Olink Biomarker | Value of 1 SD | All-cause dementia (n=504 cases) |  |  |
| --- | --- | --- | --- | --- |
|  |  | OR (95% CI) per 1 SD<sup>a</sup> | p-value per 1 SD | FDR corrected p-value<sup>b</sup> |
| ADA | 0.635 | 1.17 (1.05-1.32) | **0.0055** | **0.0098** |
| AXIN1 | 1.140 | 1.14 (1.02-1.28) | **0.0257** | **0.0352** |
| CASP-8 | 1.364 | 1.15 (1.02-1.29) | **0.0194** | **0.0279** |
| CCL3 | 1.508 | 1.14 (1.02-1.27) | **0.0259** | **0.0352** |
| CCL4 | 1.099 | 1.19 (1.06-1.33) | **0.0032** | **0.0066** |
| CCL11 | 0.697 | 1.29 (1.14-1.46) | **0.0001** | **0.0003** |
| CCL19 | 1.199 | 1.17 (1.05-1.32) | **0.0063** | **0.0108** |
| CCL20 | 1.540 | 1.18 (1.05-1.32) | **0.0041** | **0.0082** |
| CCL23 | 0.732 | 1.29 (1.14-1.46) | **<0.0001** | **0.0003** |
| CCL25 | 0.763 | 1.17 (1.03-1.32) | **0.0122** | **0.0187** |
| CCL28 | 0.548 | 1.27 (1.13-1.43) | **<0.0001** | **0.0003** |
| CD5 | 0.523 | 1.26 (1.12-1.42) | **0.0002** | **0.0007** |
| CD6 | 0.757 | 1.19 (1.05-1.34) | **0.0048** | **0.0093** |
| CD40 | 0.734 | 1.20 (1.07-1.36) | **0.0022** | **0.0050** |
| CD244 | 0.587 | 1.38 (1.22-1.57) | **<0.0001** | **0.0003** |
| CDCP1 | 0.894 | 1.20 (1.06-1.36) | **0.0030** | **0.0064** |
| CSF-1 | 0.425 | 1.24 (1.09-1.41) | **0.0013** | **0.0032** |
| CST5 | 0.698 | 1.17 (1.04-1.32) | **0.0112** | **0.0179** |
| **CX3CL1** | **0.669** | **1.41 (1.24-1.60)** | **<0.0001** | **0.0003** |
| CXCL1 | 0.901 | 1.17 (1.05-1.32) | **0.0065** | **0.0109** |
| CXCL5 | 0.957 | 1.33 (1.17-1.51) | **<0.0001** | **0.0003** |
| CXCL6 | 0.848 | 1.28 (1.14-1.44) | **0.0001** | **0.0003** |
| CXCL9 | 0.953 | 1.19 (1.05-1.34) | **0.0052** | **0.0096** |
| CXCL10 | 0.953 | 1.16 (1.03-1.30) | **0.0138** | **0.0207** |
| CXCL11 | 1.051 | 1.18 (1.05-1.33) | **0.0051** | **0.0096** |
| DNER | 0.488 | 1.36 (1.20-1.55) | **<0.0001** | **0.0003** |
| **EN-RAGE** | **1.307** | **1.41 (1.25-1.60)** | **<0.0001** | **0.0003** |
| FGF-19 | 1.089 | 1.21 (1.08-1.35) | **0.0013** | **0.0032** |
| Flt3L | 0.629 | 1.21 (1.07-1.36) | **0.0018** | **0.0042** |
| GDNF | 0.506 | 1.16 (1.03-1.30) | **0.0149** | **0.0219** |
| HGF | 0.719 | 1.34 (1.18-1.52) | **<0.0001** | **0.0003** |
| IL-7 | 0.798 | 1.14 (1.02-1.28) | **0.0246** | **0.0347** |
| IL-10 | 0.863 | 1.20 (1.07-1.35) | **0.0023** | **0.0050** |
| IL-18 | 0.763 | 1.33 (1.17-1.50) | **<0.0001** | **0.0003** |
| IL-10RA | 0.788 | 1.12 (1.01-1.25) | **0.0362** | **0.0461** |
| IL-10RB | 0.533 | 1.29 (1.14-1.46) | **0.0001** | **0.0003** |
| IL-15RA | 0.359 | 1.22 (1.09-1.38) | **0.0009** | **0.0026** |
| IL-18R1 | 0.602 | 1.27 (1.13-1.44) | **0.0001** | **0.0003** |
| LAP TGF-beta-1 | 0.574 | 1.37 (1.21-1.55) | **<0.0001** | **0.0003** |
| LIF-R | 0.503 | 1.37 (1.21-1.56) | **<0.0001** | **0.0003** |
| MCP-2 | 0.769 | 1.18 (1.05-1.33) | **0.0069** | **0.0113** |
| MCP-4 | 0.927 | 1.23 (1.08-1.39) | **0.0015** | **0.0036** |
| MMP-10 | 0.761 | 1.22 (1.08-1.37) | **0.0010** | **0.0028** |
| NT-3 | 0.544 | 1.24 (1.10-1.39) | **0.0003** | **0.0009** |
| OPG | 0.609 | 1.39 (1.22-1.58) | **<0.0001** | **0.0003** |
| PD-L1 | 0.600 | 1.30 (1.15-1.47) | **<0.0001** | **0.0003** |
| SCF | 0.624 | 1.15 (1.01-1.29) | **0.0281** | **0.0375** |
| SIRT2 | 1.157 | 1.13 (1.01-1.27) | **0.0371** | **0.0461** |
| ST1A1 | 1.304 | 1.13 (1.01-1.27) | **0.0365** | **0.0461** |
| STAMBP | 0.833 | 1.18 (1.05-1.32) | **0.0056** | **0.0098** |
| TGF-alpha | 0.829 | 1.21 (1.08-1.37) | **0.0011** | **0.0029** |
| TNFRSF9 | 0.636 | 1.23 (1.09-1.39) | **0.0006** | **0.0018** |
| TNFSF14 | 1.064 | 1.16 (1.03-1.30) | **0.0119** | **0.0186** |
| TRAIL | 0.513 | 1.31 (1.16-1.49) | **<0.0001** | **0.0003** |
| TRANCE | 0.753 | 1.14 (1.01-1.28) | **0.0344** | **0.0450** |
| TWEAK | 0.647 | 1.35 (1.19-1.53) | **<0.0001** | **0.0003** |
| VEGF-A | 0.794 | 1.40 (1.24-1.59) | **<0.0001** | **0.0003** |
| uPA | 0.604 | 1.36 (1.20-1.54) | **<0.0001** | **0.0003** |

Abbreviations: SD, standard deviation; CI, confidence interval; FDR, false discovery rate; For biomarker abbreviations, see Supplemental Table 1.
Notes: Numbers printed in bold are statistically significant ( $p < 0.05$ ). Grey shade: Independently associated with the outcome. For associations of not significantly associated biomarkers, see Supplemental Table 5.
<sup>a</sup> Multivariate logistic regression model adjusted for age (continuously), sex, education, physical activity, BMI (categorical), CVD, diabetes, depression, *APOE* genotype.
<sup>b</sup> P-values corrected for multiple testing by the Benjamini and Hochberg method.

**Table 3.** Associations of significantly associated Olink Biomarker levels with **Alzheimer’s disease** incidence.

| Olink Biomarker | Value of 1 SD | Alzheimer's disease (n=163 cases) |  |  |
| --- | --- | --- | --- | --- |
|  |  | OR (95% CI) per 1 SD<sup>a</sup> | p-value per 1 SD | FDR corrected p-value<sup>b</sup> |
| CASP-8 | 1.364 | 1.31 (1.10-1.57) | **0.0025** | **0.0156** |
| CCL23 | 0.732 | 1.43 (1.17-1.75) | **0.0004** | **0.0086** |
| CCL28 | 0.548 | 1.36 (1.14-1.61) | **0.0005** | **0.0086** |
| CD6 | 0.757 | 1.30 (1.08-1.58) | **0.0067** | **0.0254** |
| CD244 | 0.587 | 1.39 (1.14-1.70) | **0.0010** | **0.0120** |
| CX3CL1 | 0.669 | 1.35 (1.10-1.65) | **0.0034** | **0.0175** |
| CXCL5 | 0.957 | 1.37 (1.12-1.68) | **0.0023** | **0.0156** |
| CXCL6 | 0.848 | 1.34 (1.11-1.62) | **0.0026** | **0.0156** |
| DNER | 0.488 | 1.37 (1.12-1.68) | **0.0025** | **0.0156** |
| EN-RAGE | 1.307 | 1.51 (1.25-1.83) | **<0.0001** | **0.0036** |
| HGF | 0.719 | 1.36 (1.12-1.66) | **0.0017** | **0.0153** |
| IL-10RB | 0.533 | 1.33 (1.08-1.63) | **0.0066** | **0.0254** |
| LAP TGF-beta-1 | 0.574 | 1.46 (1.21-1.76) | **0.0001** | **0.0036** |
| LIF-R | 0.503 | 1.31 (1.08-1.60) | **0.0062** | **0.0254** |
| PD-L1 | 0.600 | 1.31 (1.09-1.57) | **0.0034** | **0.0175** |
| ST1A1 | 1.304 | 1.30 (1.08-1.57) | **0.0062** | **0.0254** |
| STAMBP | 0.833 | 1.26 (1.06-1.51) | **0.0108** | **0.0370** |
| TGF-alpha | 0.829 | 1.26 (1.05-1.52) | **0.0140** | **0.0458** |
| TRAIL | 0.513 | 1.30 (1.06-1.59) | **0.0104** | **0.0370** |
| TWEAK | 0.647 | 1.38 (1.13-1.69) | **0.0016** | **0.0153** |
| VEGF-A | 0.794 | 1.32 (1.09-1.60) | **0.0042** | **0.0202** |
| uPA | 0.604 | 1.40 (1.16-1.71) | **0.0006** | **0.0086** |

Abbreviations: SD, standard deviation; CI, confidence interval; FDR, false discovery rate; For biomarker abbreviations, see Supplemental Table 1.
Notes: Numbers printed in bold are statistically significant ( $p < 0.05$ ). Grey shade: Independently associated with the outcome. For associations of not significantly associated biomarkers, see Supplemental Table 6.
<sup>a</sup> Multivariate logistic regression model adjusted for age (continuously), sex, education, physical activity, BMI (categorical), CVD, diabetes, depression, *APOE* genotype.
<sup>b</sup> P-values corrected for multiple testing by the Benjamini and Hochberg method.

**Table 4.**
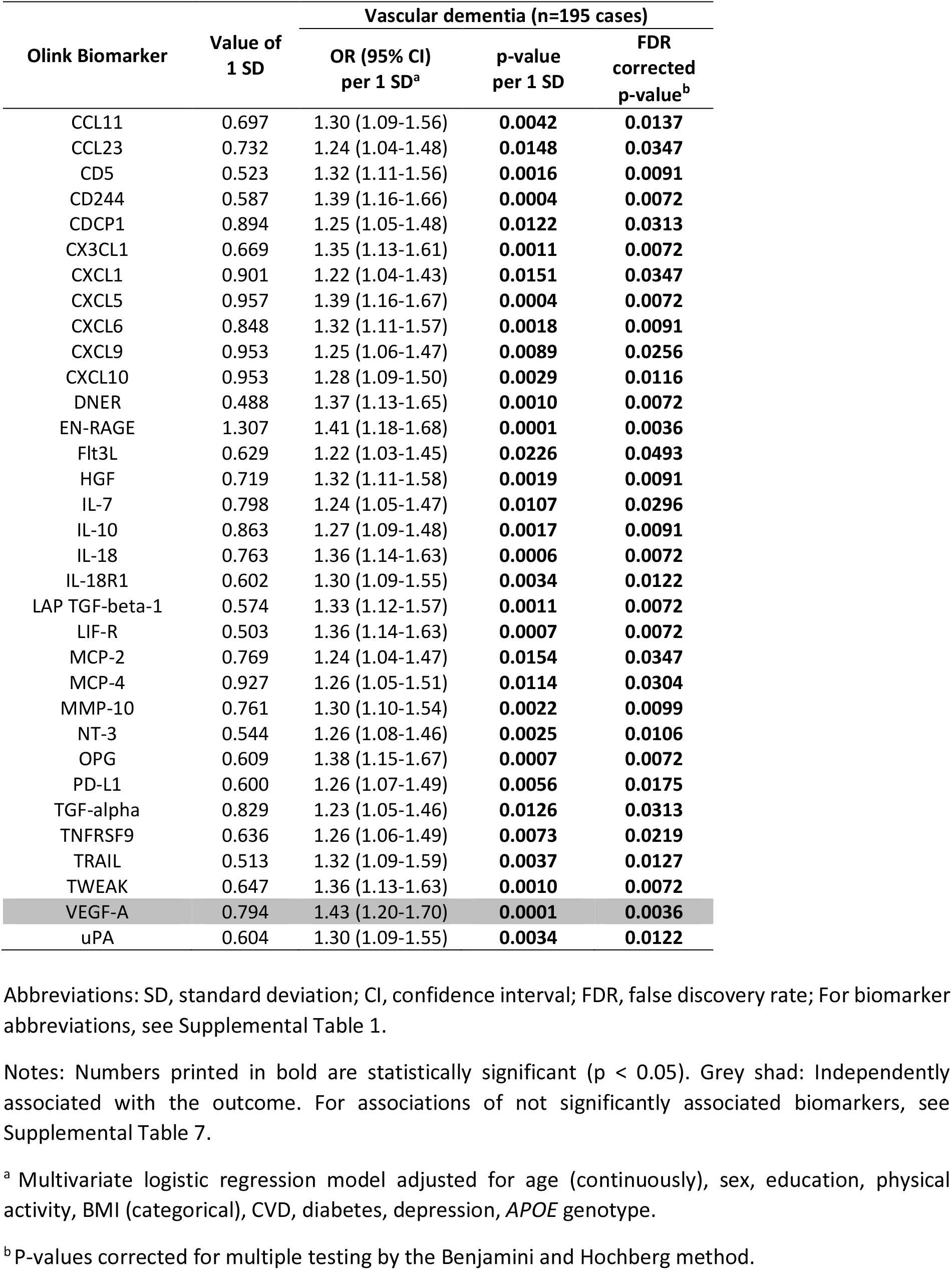
Associations of significantly associated Olink Biomarker levels with **vascular dementia** incidence.

When the two independent biomarkers for all-cause dementia were added simultaneously to the logistic regression models, the OR point estimates per 1 SD increase were attenuated but remained statistically significant (CX3CL1, OR [95% CI]: 1.29 [1.13-1.47], p=0.0002; EN-RAGE, OR [95% CI]: 1.31 [1.15-1.49], p<0.0001). This was also the case for the two independent biomarkers for AD (EN-RAGE, OR [95% CI]: 1.37 [1.10-1.68], p =0.0048; LAP TGF-beta-1, OR [95% CI]: 1.28 [1.04-1.58], p=0.0187). For VD, only one independent biomarker was included (VEGF-A, OR [95% CI]: 1.43 [1.20-1.70], p<0.0001). The dose-response curves of these five biomarker-dementia outcome associations are shown in Figure 1. The risk of VD seems to start to increase only at higher VEGF-A levels (> 60^th^ percentile). The other four biomarker-dementia associations show a more or less linear risk increase over the whole biomarker level distribution.

**Figure 1.**
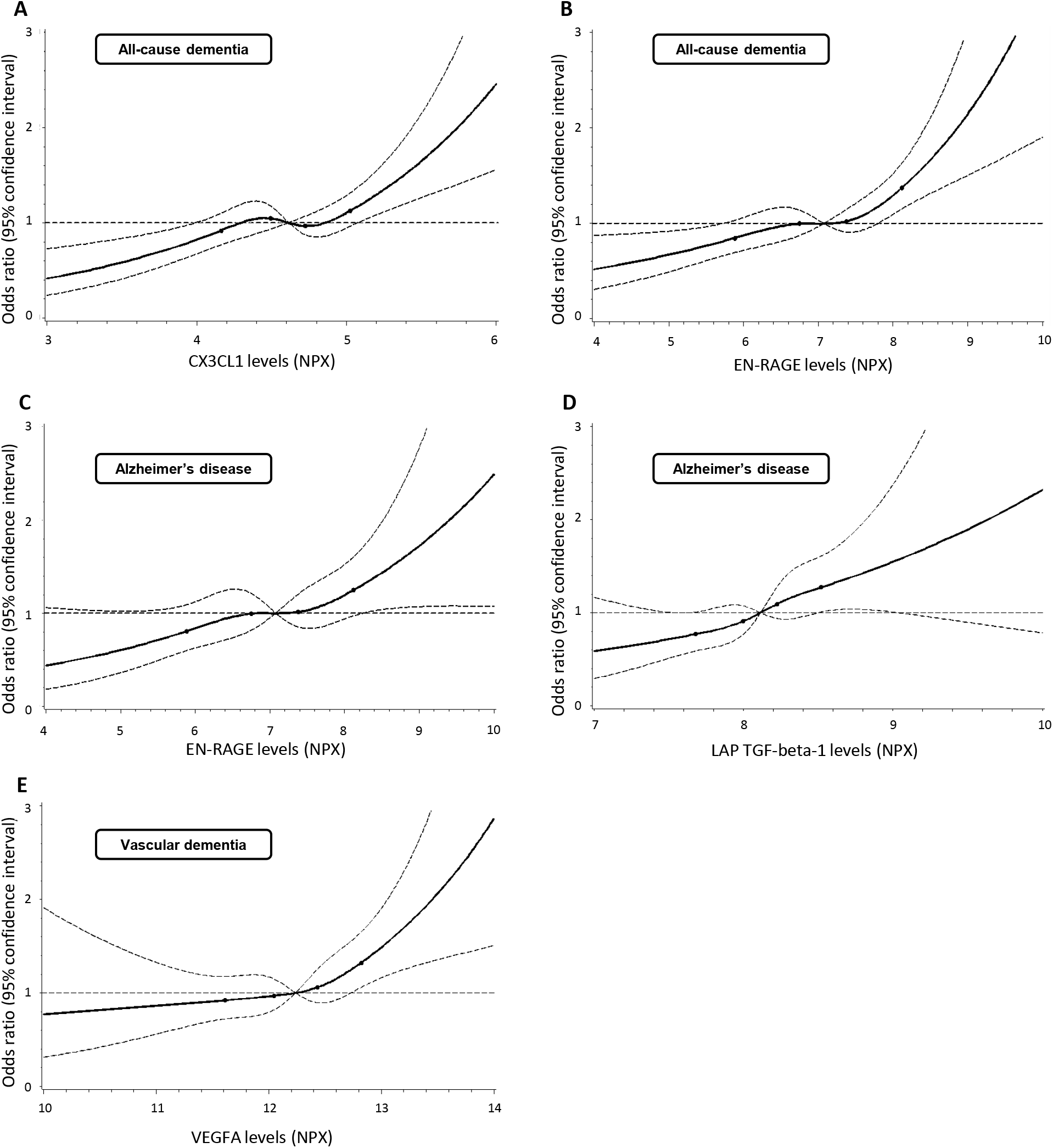
Association of all-cause dementia with (A) CX3CL1 and (B) EN-RAGE, Alzheimer’s disease with (C) EN-RAGE and (D) LAP TGF-beta-1, and vascular dementia with (E) VEGFA in a spline regression model adjusted for age (continuously), sex, education, physical activity, BMI (categorical), cardiovascular disease, diabetes, depression, APOE genotype. Solid lines: estimation; dashed curved lines: 95% confidence interval limits; dashed horizontal line: reference line (hazard ratio = 1); dots: knots (20^th^, 40^th^, 60^th^, and 80^th^ percentile). Abbreviations: NPX, Normalized Protein eXpression

The results for these five selected biomarker-dementia endpoint associations are shown stratified for age, sex, obesity, diabetes, CVD, *APOE* ε4 in Supplemental Tables 12-16. Generally, results were similar in subgroups defined by the first four factors. For *APOE* ε4, there was a consistent pattern towards stronger associations of inflammation biomarkers among *APOE* ε4 negative subjects. In line with this observation, the only statistically significant interaction found was between *APOE* ε4 polymorphism and the biomarker EN-RAGE for all-cause dementia (p=0.024, Supplemental Table 13).

The results of the sensitivity analyses are also shown in Supplemental Tables 12-16. When stratified by time of diagnosis, all selected biomarkers had a stronger association with dementia diagnoses occurring in the first ten years of follow-up. However, significant associations were also observed for diagnoses in later years of follow-up. Besides, excluding subjects who died before their 80^th^ birthday or had a sign of acute infection (CRP level >20mg/L) did not alter the results to any relevant extent.

## 4. Discussion

To our knowledge, this is the first prospective cohort study to analyze inflammation-related, blood-based biomarkers for all-cause dementia, AD, and VD incidence. We identified a high number of statistically significantly associated proteins with at least one of the outcomes, even after FDR correction. However, only a few biomarkers were strongly and independently associated with dementia outcomes because of a high inter-correlation between the biomarkers. The identified independent biomarkers include CX3CL1 (associated with all-cause dementia), EN-RAGE (associated with all-cause dementia and AD), LAP TGF-beta-1 (associated with AD), and VEGF-A (associated with VD). Each of these biomarkers is only one marker of an inflammatory protein cluster, in which the majority of biomarkers are associated with dementia.

### 4.1. Previous studies examining a set of inflammatory biomarkers

A few previous studies, mostly with a cross-sectional study design, investigated the association between single inflammatory biomarkers and all-cause dementia or AD^6–9^. To our knowledge, only one previous study examined a whole panel of inflammatory biomarkers for dementia as the outcome in a cross-sectional design. In the BioFINDER study, Whelan and colleagues measured 270 proteins with the Olink immunoassay in cerebrospinal fluid (CSF) and plasma of 161 AD patients, 75 amyloid beta positive (Aβ+) patients with mild cognitive impairment (MCI+), and 415 amyloid beta negative (Aβ-) cognitively normal individuals (MCI-)^25^. Interestingly, approximately half of the CSF proteins correlated at least modestly with their analogues in plasma, indicating that findings in plasma samples partially reflected the situation in CSF. Compared to Aβ-/MCI-individuals, CSF levels of 32 proteins and plasma levels of 33 proteins were statistically significantly associated with AD (False discovery corrected p-value < 0.05). The comparison of Aβ+/MCI+ patients with Aβ-/MCI-individuals, were replicated in an independent cohort. Thereby, 10 CSF and six plasma markers could be replicated. However, a replication analysis in an independent sample was not performed for AD. Six of the identified 33 proteins for AD in plasma samples corresponded with our findings in serum samples and can now be considered replicated by our study (Casp-8, CXCL5, CXCL6, ST1A1, TRAIL, uPA).

However, the study of Whelan and colleagues was cross-sectional and evidence from longitudinal studies on this field is still sparse^10^. The recently published longitudinal study of Walker et al. reported statistically significant associations between inflammatory biomarkers measured in midlife (C-reactive protein and a composite score of fibrinogen, white blood cell count, von Willebrand factor, and factor VIII) and cognitive decline over 20 years in a population-based cohort study with 12,336 participants^26^. Our longitudinal results with a broad panel of inflammatory proteins and the endpoints all-cause dementia, AD and VD complement and expand these findings.

### 4.2. Independently associated biomarkers

The independently associated biomarkers CX3CL1, EN-RAGE, LAP TGF-beta-1 and VEGF-A likely play various important roles in the pathogenesis of dementia. While CX3CL1 is discussed to regulate microglial activation^27^ and LAP TGF-beta-1 to be protective against Aß deposition^13, 28^, VEGF-A is thought to increase the permeability of the blood brain barrier^29^. EN-RAGE is less studied in the context of dementia^30^. This was the first longitudinal cohort study, which reported an association of EN-RAGE with all-cause dementia and AD. More details about plausible mechanisms of these four biomarkers and other prominently discussed inflammatory proteins in the development of dementia are discussed in **Supplemental Text 3**. Furthermore, we discuss the *APOE* ε4 specific results of our study there.

### 4.3. Strengths and limitations

The strengths of this study comprise a large sample size, the representative sample of an older adult population, a long follow-up period (17 years), and the prospective cohort design. Moreover, the diversity of inflammatory biomarkers (72 biomarkers analyzed) and the high sensitivity and specificity of Olink’s proximity extension assays^31, 32^ used for the biomarker measurements can be assigned to the study’s strengths.

The observational study design is one of the limitations of this study. Although analyses were controlled for confounders, residual confounding cannot be entirely excluded. Apart from this, the latency between the onset and the clinical diagnosis of dementia can be longer than the follow-up time of 17 years^33^. However, results were still statistically significant after excluding events in the first ten years of follow-up. Thus, we can assume that there is no strong indication of reverse causality in our study results.

In the ESTHER study, dementia information is collected via GPs. After a referral to various neurologists, psychiatrists, memory clinics, or other specialized providers in the study region, diagnoses were obtained from the medical records of specialists. This process reflects the community-based clinical setting in Germany. Therefore, dementia diagnostics were performed heterogeneously, and dementia subtypes were often not assessed. This may be one reason why the ratio of AD to VD diagnoses is not as high in our study as in other studies with homogenous subtype diagnostics based on biomarkers of AD pathology among all study participants.

Due to cost issues, biomarker measurements were only performed for baseline blood samples and could not be repeated in follow-ups. Furthermore, findings could not be replicated in another independent study, which should be aimed at future research.

The biomarkers TNF and IFN-gamma had to be excluded since the proportion of values below LOD was > 25% in the total study sample. After Olink improved the inflammation panel in 2019, better results could be achieved for these biomarkers. However, the improved panel was only used in a fraction of our study sample (n = 440). When analyzing the data only in this sub-sample, TNF and IFN-gamma showed significant associations for all three analyzed dementia outcomes.

Lastly, it has to be stated that our study results refer to an almost exclusively Caucasian population with blood samples taken between the ages of 50 and 75 years and may not be generalized to other types of populations.

### 4.4. Conclusion

This study showed that 58 out of 72 tested proteins of the inflammatory proteome in blood were significantly associated with all-cause dementia incidence even after correction for multiple testing. Several inflammatory proteins were further associated with AD and VD. The biomarkers CX3CL1, EN-RAGE, LAP TGF-beta-1, and VEGF-A, had strong and independent associations with dementia outcomes and may have great potential as drug targets, early diagnostic markers, and components of dementia prediction scores. However, due to the observed high inter-correlation of inflammatory biomarkers, it should be noted that not only single biomarkers but also clusters of increased inflammatory protein levels may play a role in dementia pathogenesis or risk prediction. The complex interrelationships in these clusters are not yet understood and require further research.

## Data Availability

Data are only accessible on request

## Acknowledgements

The authors have no conflicts of interest to disclose. Financial support for research staff involved in this project was granted by the Baden-Württemberg State Ministry of Science, Research and Arts (Stuttgart, Germany), the Robert-Bosch-Stiftung (Stuttgart, Germany) and the Klaus-Tschira-Stiftung gGmbH (Heidelberg, Germany). The ESTHER study was funded by grants from the Baden-Württemberg state Ministry of Science, Research and Arts (Stuttgart, Germany), the Federal Ministry of Education and Research (Berlin, Germany), the Federal Ministry of Family Affairs, Senior Citizens, Women and Youth (Berlin, Germany), and the Saarland state ministry for Social Affairs, Health, Women and Family Affairs (Saarbrücken, Germany).

## Supplemental Material

### Supplemental Text 1

#### Biomarker assessment

Inflammation-related, blood-based proteins were measured from serum samples collected during the health checkup at baseline (2000-2002). Blood samples were sent to the study centre and stored at −80°C until biomarker measurements took place in March 2018, December 2018, and September 2020 (referred to as time points t1, t2 and t3 in the following). At the time of the measurements, 10-25 µl of serum was extracted from different aliquots that have been thawed twice and sent with dry ice to the laboratories, which analyzed the samples with the Olink Target 96 Inflammation panel, Olink Proteomics, Uppsala, Sweden. At t1 and t2, samples were analyzed in the laboratory of Olink Proteomics, Uppsala Science Park, SE-75183 Uppsala, Sweden. At t3, the measurements were performed in the Research Unit Protein Science, German Research Center for Environmental Health, Helmholtz Center Munich, Heidemannstraße 1, 80939 München, Germany.

The Olink panels are based on a Proximity Extension Assay technology (PEA)^1, 2^. Details on the reliability and stability of the technology are described elsewhere^3^. In brief, oligonucleotide labelled antibody probe pairs are allowed to bind to their respective target proteins in the samples. Only if two antibodies are in close proximity, a polymerase chain reaction (PCR) reporter sequence is formed by DNA polymerization. This sequence is detected and quantified using high throughput real-time quantitative PCR (qPCR) (Fluidigm® Biomark^TM^ HD system). The Olink Target 96 Inflammation panel allows the measurement of 92 biomarkers per sample. A list of all biomarkers of this panel is displayed in Supplemental Table 1.

At t1, t2 and t3, 22, 15 and 5 plates were used, respectively. To avoid batch effects, cases and controls were randomly distributed across plates and adjusted according to included interpolate controls. The average intra-assay coefficient of variance among all 92 measured biomarkers was 7%, 4% and 3% at t1, t2 and t3, respectively. The average inter-assay coefficient of variance was 12%, 10% and 10% at t1, t2 and t3, respectively. Furthermore, the quality of each serum samples was assessed by Olink technology. All samples were measured successfully, and the number of quality control warnings was below 4% in all three timepoints. Of the 1,435 randomly selected controls and 393 incident dementia cases, 46 serum samples of participants were excluded due to a quality control warning by Olink. Protein levels are reported as Normalized Protein eXpression (NPX) values, a relative quantification unit logarithmically related to protein concentration. The number of samples with values below the lower limit of detection (LOD) varied strongly by biomarker and is shown in Supplemental Table 1 (Appendix). In total, 20 biomarkers with > 25 % of the values below LOD were excluded from all analyses (grey shaded biomarkers in Supplemental Table 1). Thereby, 72 out of the 92 biomarkers were considered evaluable markers. Biomarker values below the LOD were replaced by LOD/√2. The normalization of raw data was conducted with the R (R Core Team, 2020, version 3.6.3) package “OlinkAnalyze”, developed and maintained by the Olink Proteomics Data Science Team^4^. For this procedure, bridging samples were used to normalize data from three different measurement time points.

#### Covariate assessment

Data on sex, age, education, body mass index (BMI), physical activity, and lifetime history of depression were collected during baseline assessment through a standardized self-administered questionnaire. The history of coronary heart disease (CHD) and diabetes mellitus were obtained from physician diagnoses. Furthermore, anti-diabetic drugs reported by the GP were used to complement diabetes mellitus diagnoses. Participants were considered to have cardiovascular disease (CVD) based upon CHD diagnoses from GPs or self-reported history of myocardial infarction, stroke, pulmonary embolism, or revascularization of coronary arteries. TaqMan single-nucleotide polymorphism (SNP) genotyping assays were used to determine Apolipoprotein E (*APOE*) genotypes. More precisely, genotypes were analyzed in an endpoint allelic discrimination using a PRISM 7000 Sequence detection system (Applied Biosystems)^5^.

### Supplemental Text 2

#### Sensitivity analyses

Several sensitivity analyses were performed. To check for potential reverse causality, the associations between the independent biomarkers and dementia endpoints were analyzed stratified by time of diagnosis (in the first ten years of follow-up vs later years). Competing risk of death was examined by excluding subjects without dementia diagnosis who died before their 80^th^ birthday, the average life expectancy of the cohort’s population. Fractional polynomials with first-order terms were used to determine each biomarker’s best fitting function with the outcomes^6^. Since the linear function was the best fitting one for almost all biomarkers (68 of 72), all were modelled linearly. Finally, to examine the impact of persons with a potential acute infection on the overall results, subjects with C-reactive protein (CRP) levels > 20 mg/L were excluded.

#### Imputation of missing covariate values

To our knowledge, missing values of covariates were missing at random. The highest proportion of missing values was found for *APOE* polymorphism (7.7%). Thus, multiple imputation was used to impute missing values. Variables shown in Table 1 were used for the imputation model. Twenty data sets were imputed with the Markov Chain Monte Carlo (MCMC) method separately by sex with the SAS procedure PROC MI. All analyses were performed based on those 20 datasets with the SAS procedure PROC MIANALYZE.

Statistical tests were two-sided using an alpha level of 0.05. All statistical analyses were conducted with the Statistical Analysis System (SAS, version 9.4, Cary, North Carolina, USA).

### Supplemental Text 3

**Previous studies and pathways to dementia of selected biomarkers with independent associations**

#### CX3CL1

The biomarker CX3CL1, which is also commonly known as Fractalkine in humans, was independently associated with all-cause dementia and among the list of statistically significant biomarkers for AD and VD in our study. This biomarker is a chemokine binding to its receptor C-X3-C motif chemokine receptor 1 (CX3CR1) in a one-to-one relationship. While CX3CL1 is usually expressed in neurons, CX3CR1 is expressed on microglia. In the case of neuroinflammation, CX3CL1 regulates microglial activation by reducing the release of pro-inflammatory products^7^. Whether the effect of CX3CL1 is neuroprotective or neurotoxic in diseases like dementia is still controversially discussed in the literature. The current opinion is that this depends on the disease state, the affected CNS area, and the local concentration of the CX3CL1/CX3CR1 complex^7, 8^.

Nevertheless, due to the regulatory function in inflammation, this biomarker is a promising therapeutic target. A recent Polish study reported on the predictive ability of CX3CL1 as a biomarker in the early development of mild cognitive impairment (MCI) and AD^9^. In this study, significantly higher CSF and blood levels of CX3CL1 were found in MCI and AD patients compared to cognitively healthy controls. We now confirm these results with longitudinal data, including 17 years of follow-up.

#### EN-RAGE

EN-RAGE was independently associated with all-cause dementia and AD. In addition, EN-RAGE was significantly associated with VD. EN-RAGE is also often referenced as S100-A12. The S100-protein family has already been shown multiple times to be related to AD^10^. However, the S100-A12 protein (EN-RAGE) is the least studied S100 protein in the context of AD and dementia^10^. In the only available study, Shepherd and colleagues revealed associations of EN-RAGE with senile plaques, reactive glia, and neurons in brain samples of sporadic and familial (PS-1) AD cases in a cross-sectional study^11^. Our study is the first longitudinal cohort study reporting on this association.

EN-RAGE is a calcium-, zinc-, and copper-binding protein. In previous studies, it was shown to be associated with diseases like heart failure^12^ and coronary artery disease (CAD) in diabetes patients^13^. Recently, Feng and colleagues reported significantly elevated EN-RAGE concentrations in patients with traumatic brain injury compared to controls^14^. In this study, EN-RAGE showed great potential as a marker for ongoing inflammatory processes in the brain. RAGE, the receptor EN-RAGE binds to, is additionally known to be involved in inflammatory processes related to ageing and neurodegeneration^15, 16^.

#### LAP-TGF-beta-1

LAP TGF-beta-1 is an anti-inflammatory cytokine that was independently associated with AD in our study (OR [95% CI]: 1.46 [1.21-1.76]). Additionally, it was significantly associated with all-cause dementia and VD. This biomarker consists of two components, latency-associated peptide (LAP) and transforming growth factor beta-1 (TGF-beta-1), which are non-covalently linked to each other in the intracellular environment. Thereby, LAP keeps TGF-beta-1 biologically inactive^17^. When activated, TGF-beta-1 binds to its receptor transforming growth factor-ß receptor type I (TßR-1), protecting neurons against Aß deposits and apoptosis^18, 19^. However, controversial findings have been reported on concentrations of this biomarker in AD^20^. The current theory is that the level of TGF-beta-1 in the body might depend on disease progression^20^. According to this theory, the reported elevated levels of TGF-beta-1 in our study might show an early response to commencing neurodegenerative processes in AD. A recent study additionally reported on the specificity of TGF-beta-1 for AD and VD compared to Parkinson’s disease dementia (PDD)^21^.

#### VEGF-A

In our cohort, VEGF-A was independently associated with VD (OR [95% CI]: 1.43 [1.20-1.70]) and also significantly associated with all-cause dementia and AD. VEGF-A belongs to the vascular endothelial growth factor (VEGF) family and induces endothelial cell growth, cell migration, and permeabilization of blood vessels. Like other members of this family, VEGF-A induces the receptors VEGF receptor 1 and 2 (VEGFR-1 and VEGFR-2)^22^. In VD, VEGF-A is reported to be involved in microvessel loss and blood-brain barrier breakdown^23^. It was shown in the same study in mice that VEGF-A is involved in the hypoxia-inducible factor 1α-Lipocalin2-VEGFA (HIF-1α-LCN2-VEGFA) axis. Other groups have also shown an involvement of VEGF-A in increasing blood-brain barrier permeability^24, 25^. Hence, blocking VEGF-A signalling might be a promising therapeutic target^26^.

#### Inflammatory proteins prominently discussed in dementia research and biomarker clusters

Interestingly, the frequently discussed inflammatory biomarker IL-6 was not significantly associated with any dementia outcome in our study but highly correlated with EN-RAGE and VEGF-A [6-8]. However, apart from the inflammatory biomarkers discussed above, many others were statistically significantly associated with dementia outcomes as well but highly correlated with the highlighted proteins. IL-10, for example, is currently discussed by others as a risk factor for AD^27^. In our study, IL-10 was also statistically significantly associated with all-cause dementia and VD even after correction for multiple testing. In addition, a subunit of the IL-10 receptor (IL-10RB) was significantly associated with all-cause dementia and AD. Both IL-10 and IL-10RB were highly correlated with VEGF-A and IL-10RB, additionally with LAP TGF-beta-1 and CX3CL1. Due to the high correlation of these biomarkers, it is not possible to decide with our study design which of the biomarkers are the most clinically relevant ones and are causally associated with the outcome. Basic research is needed to elucidate this open question and the role of the identified biomarkers in the etiology of dementia. The underlying mechanisms are likely to be complex because it is known that the multifactorial process of inflammation comes along with increases in the levels of many inflammatory proteins. Therefore, it might be necessary to look into inflammatory protein networks/clusters rather than focusing on single proteins in future studies. For example, for AD, we identified two such protein clusters. The EN-RAGE and the LAP TGF-beta-1 cluster consist of respectively nine inflammatory proteins significantly associated with AD (Supplemental Tables 9 and 10). The overlap of the two clusters is only three proteins (HGF, CD244 and uPA).

#### Role of *APOE* ε4 polymorphism

*APOE* ε4 negative subjects had stronger associations between inflammation biomarkers and dementia outcomes than *APOE* ε4 positive individuals. Interestingly, the interaction of *APOE* ε4 and EN-RAGE for all-cause dementia was statistically significant. One explanation could be that the absolute AD risk of *APOE* ε4 carriers is so pronounced that the additional presence or absence of a weaker risk factor, such as inflammation, may not have much impact. In contrast, the potential impact of inflammation on the total dementia risk of *APOE* ε4 non-carriers is relatively high compared to other dementia risk factors.

---

**Supplemental Figure 1.**
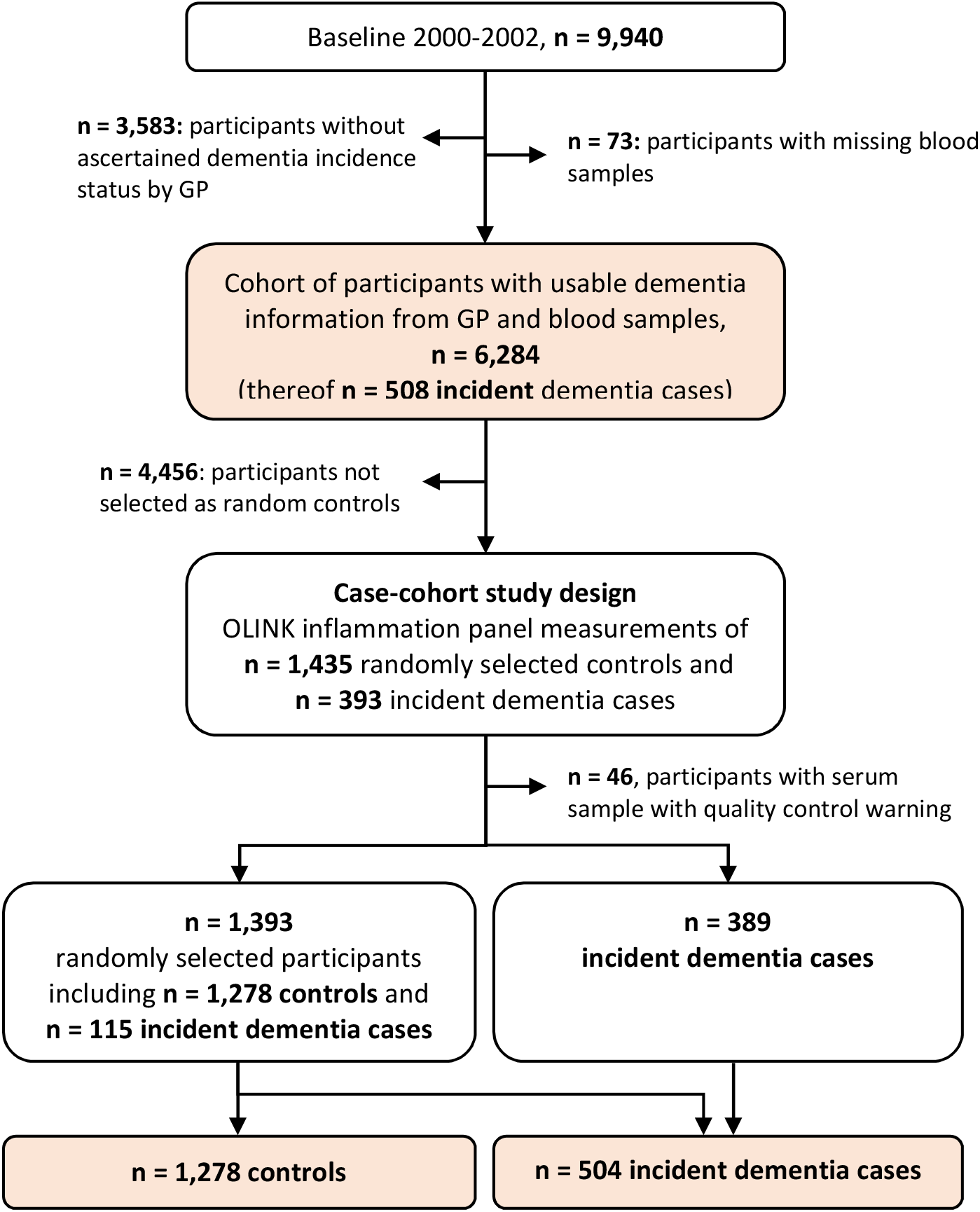
Flowchart of dementia ascertainment during the 14- and 17-year follow-up of the ESTHER study and selection of the study population for this research project. **Abbreviations:** GP, general practitioner.

**Supplemental Table 1.**
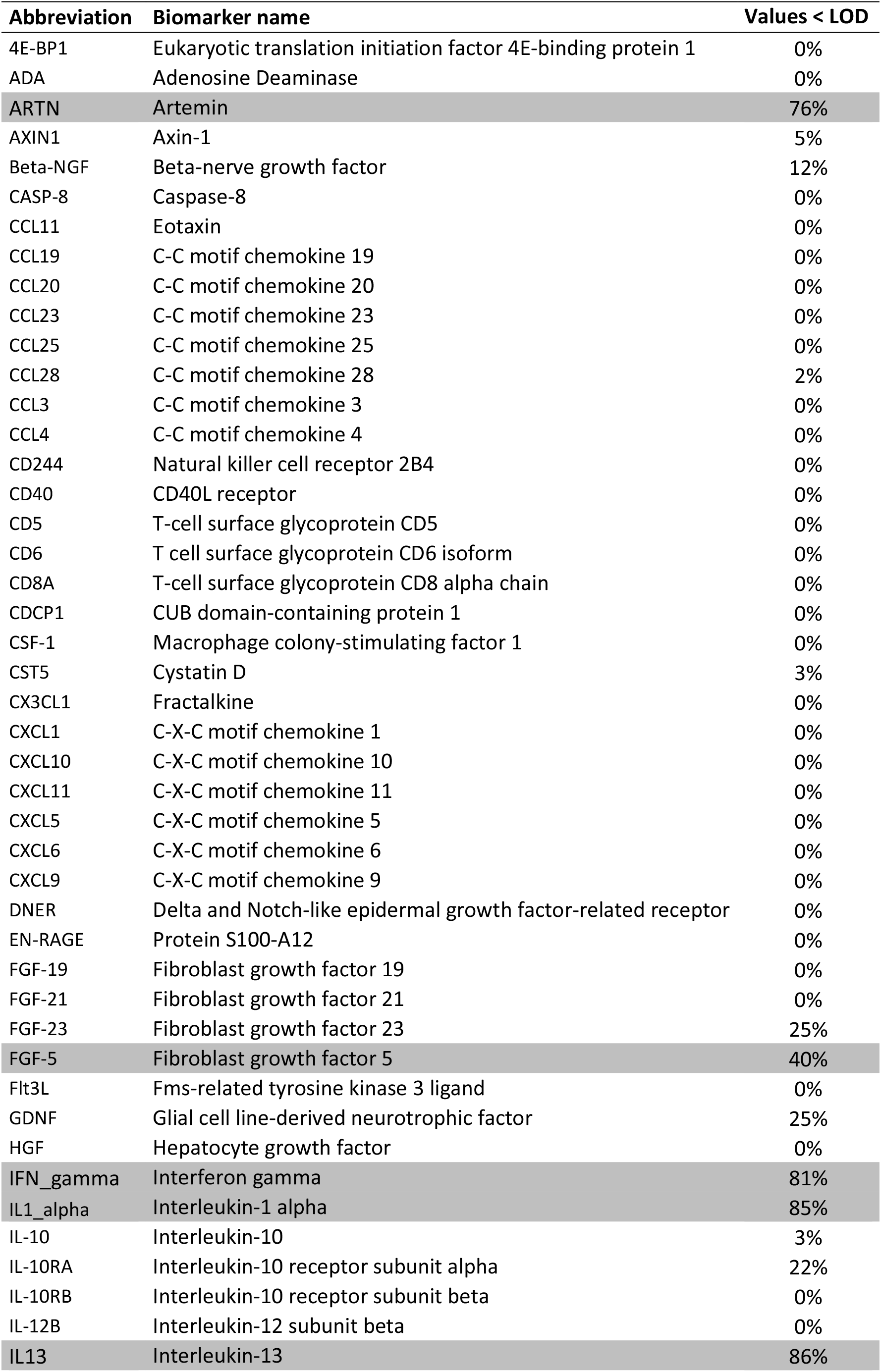
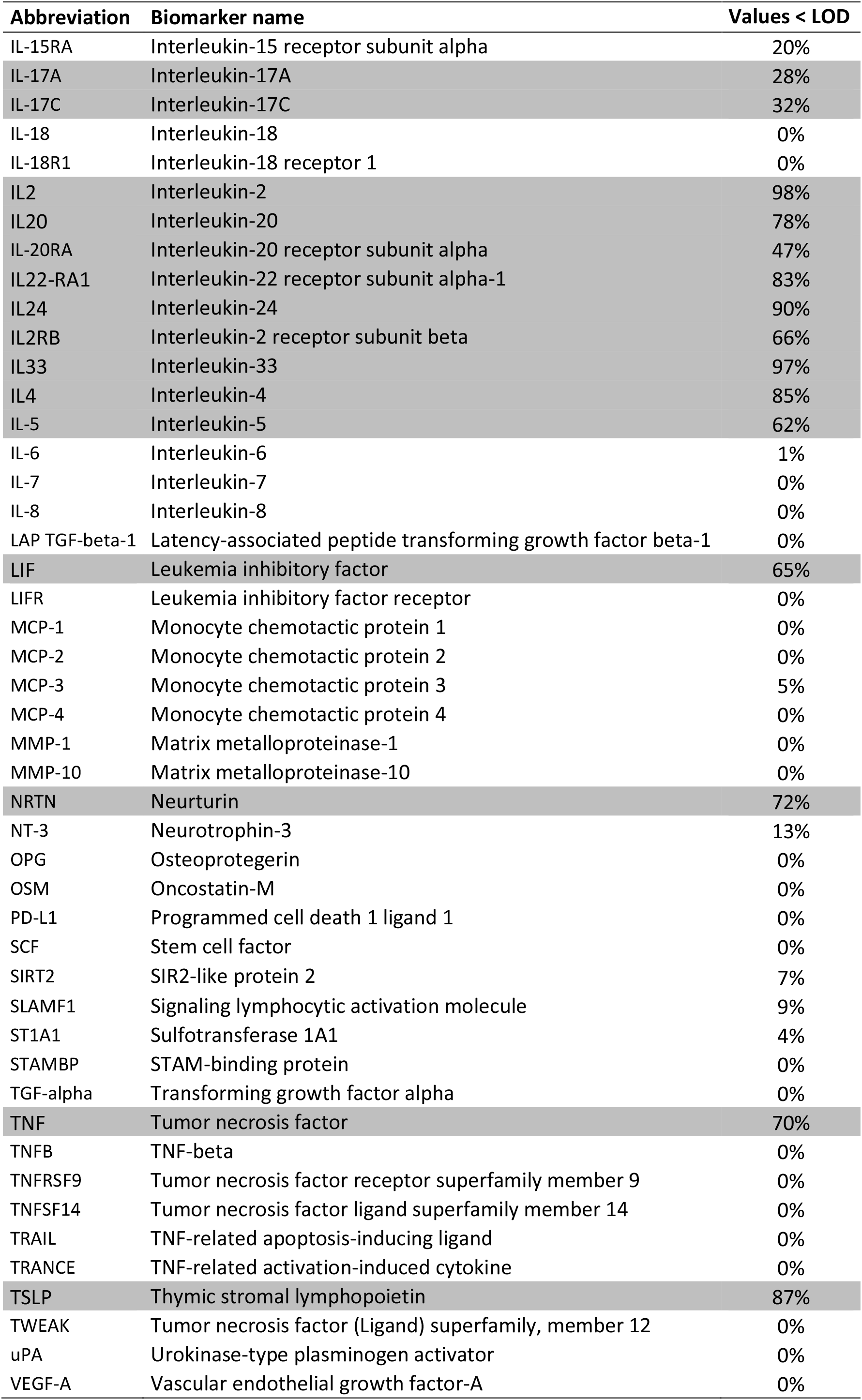
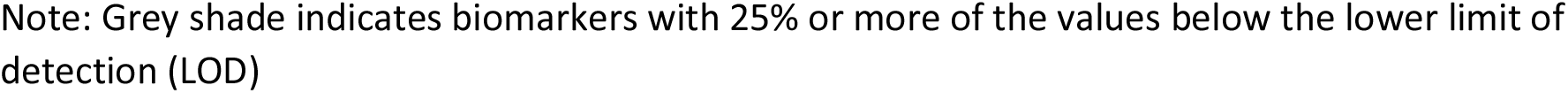
List of biomarkers measured with Olink Proseek® Multiplex Inflammation I^96x96^ kits.

| Abbreviation | Biomarker name | Values < LOD |
| --- | --- | --- |
| 4E-BP1 | Eukaryotic translation initiation factor 4E-binding protein 1 | 0% |
| ADA | Adenosine Deaminase | 0% |
| **ARTN** | **Artemin** | **76%** |
| AXIN1 | Axin-1 | 5% |
| Beta-NGF | Beta-nerve growth factor | 12% |
| CASP-8 | Caspase-8 | 0% |
| CCL11 | Eotaxin | 0% |
| CCL19 | C-C motif chemokine 19 | 0% |
| CCL20 | C-C motif chemokine 20 | 0% |
| CCL23 | C-C motif chemokine 23 | 0% |
| CCL25 | C-C motif chemokine 25 | 0% |
| CCL28 | C-C motif chemokine 28 | 2% |
| CCL3 | C-C motif chemokine 3 | 0% |
| CCL4 | C-C motif chemokine 4 | 0% |
| CD244 | Natural killer cell receptor 2B4 | 0% |
| CD40 | CD40L receptor | 0% |
| CD5 | T-cell surface glycoprotein CD5 | 0% |
| CD6 | T cell surface glycoprotein CD6 isoform | 0% |
| CD8A | T-cell surface glycoprotein CD8 alpha chain | 0% |
| CDCP1 | CUB domain-containing protein 1 | 0% |
| CSF-1 | Macrophage colony-stimulating factor 1 | 0% |
| CST5 | Cystatin D | 3% |
| CX3CL1 | Fractalkine | 0% |
| CXCL1 | C-X-C motif chemokine 1 | 0% |
| CXCL10 | C-X-C motif chemokine 10 | 0% |
| CXCL11 | C-X-C motif chemokine 11 | 0% |
| CXCL5 | C-X-C motif chemokine 5 | 0% |
| CXCL6 | C-X-C motif chemokine 6 | 0% |
| CXCL9 | C-X-C motif chemokine 9 | 0% |
| DNER | Delta and Notch-like epidermal growth factor-related receptor | 0% |
| EN-RAGE | Protein S100-A12 | 0% |
| FGF-19 | Fibroblast growth factor 19 | 0% |
| FGF-21 | Fibroblast growth factor 21 | 0% |
| FGF-23 | Fibroblast growth factor 23 | 25% |
| **FGF-5** | **Fibroblast growth factor 5** | **40%** |
| Flt3L | Fms-related tyrosine kinase 3 ligand | 0% |
| GDNF | Glial cell line-derived neurotrophic factor | 25% |
| HGF | Hepatocyte growth factor | 0% |
| **IFN_gamma** | **Interferon gamma** | **81%** |
| **IL1_alpha** | **Interleukin-1 alpha** | **85%** |
| IL-10 | Interleukin-10 | 3% |
| IL-10RA | Interleukin-10 receptor subunit alpha | 22% |
| IL-10RB | Interleukin-10 receptor subunit beta | 0% |
| IL-12B | Interleukin-12 subunit beta | 0% |
| **IL13** | **Interleukin-13** | **86%** |

| **Abbreviation** | **Biomarker name** | **Values < LOD** |
| --- | --- | --- |
| IL-15RA | Interleukin-15 receptor subunit alpha | 20% |
| IL-17A | Interleukin-17A | 28% |
| IL-17C | Interleukin-17C | 32% |
| IL-18 | Interleukin-18 | 0% |
| IL-18R1 | Interleukin-18 receptor 1 | 0% |
| IL2 | Interleukin-2 | 98% |
| IL20 | Interleukin-20 | 78% |
| IL-20RA | Interleukin-20 receptor subunit alpha | 47% |
| IL22-RA1 | Interleukin-22 receptor subunit alpha-1 | 83% |
| IL24 | Interleukin-24 | 90% |
| IL2RB | Interleukin-2 receptor subunit beta | 66% |
| IL33 | Interleukin-33 | 97% |
| IL4 | Interleukin-4 | 85% |
| IL-5 | Interleukin-5 | 62% |
| IL-6 | Interleukin-6 | 1% |
| IL-7 | Interleukin-7 | 0% |
| IL-8 | Interleukin-8 | 0% |
| LAP TGF-beta-1 | Latency-associated peptide transforming growth factor beta-1 | 0% |
| LIF | Leukemia inhibitory factor | 65% |
| LIFR | Leukemia inhibitory factor receptor | 0% |
| MCP-1 | Monocyte chemotactic protein 1 | 0% |
| MCP-2 | Monocyte chemotactic protein 2 | 0% |
| MCP-3 | Monocyte chemotactic protein 3 | 5% |
| MCP-4 | Monocyte chemotactic protein 4 | 0% |
| MMP-1 | Matrix metalloproteinase-1 | 0% |
| MMP-10 | Matrix metalloproteinase-10 | 0% |
| NRTN | Neurturin | 72% |
| NT-3 | Neurotrophin-3 | 13% |
| OPG | Osteoprotegerin | 0% |
| OSM | Oncostatin-M | 0% |
| PD-L1 | Programmed cell death 1 ligand 1 | 0% |
| SCF | Stem cell factor | 0% |
| SIRT2 | SIR2-like protein 2 | 7% |
| SLAMF1 | Signaling lymphocytic activation molecule | 9% |
| ST1A1 | Sulfotransferase 1A1 | 4% |
| STAMBP | STAM-binding protein | 0% |
| TGF-alpha | Transforming growth factor alpha | 0% |
| TNF | Tumor necrosis factor | 70% |
| TNFB | TNF-beta | 0% |
| TNFRSF9 | Tumor necrosis factor receptor superfamily member 9 | 0% |
| TNFSF14 | Tumor necrosis factor ligand superfamily member 14 | 0% |
| TRAIL | TNF-related apoptosis-inducing ligand | 0% |
| TRANCE | TNF-related activation-induced cytokine | 0% |
| TSLP | Thymic stromal lymphopoietin | 87% |
| TWEAK | Tumor necrosis factor (Ligand) superfamily, member 12 | 0% |
| uPA | Urokinase-type plasminogen activator | 0% |
| VEGF-A | Vascular endothelial growth factor-A | 0% |

Note: Grey shade indicates biomarkers with 25% or more of the values below the lower limit of detection (LOD)

**Supplemental Table 2.**
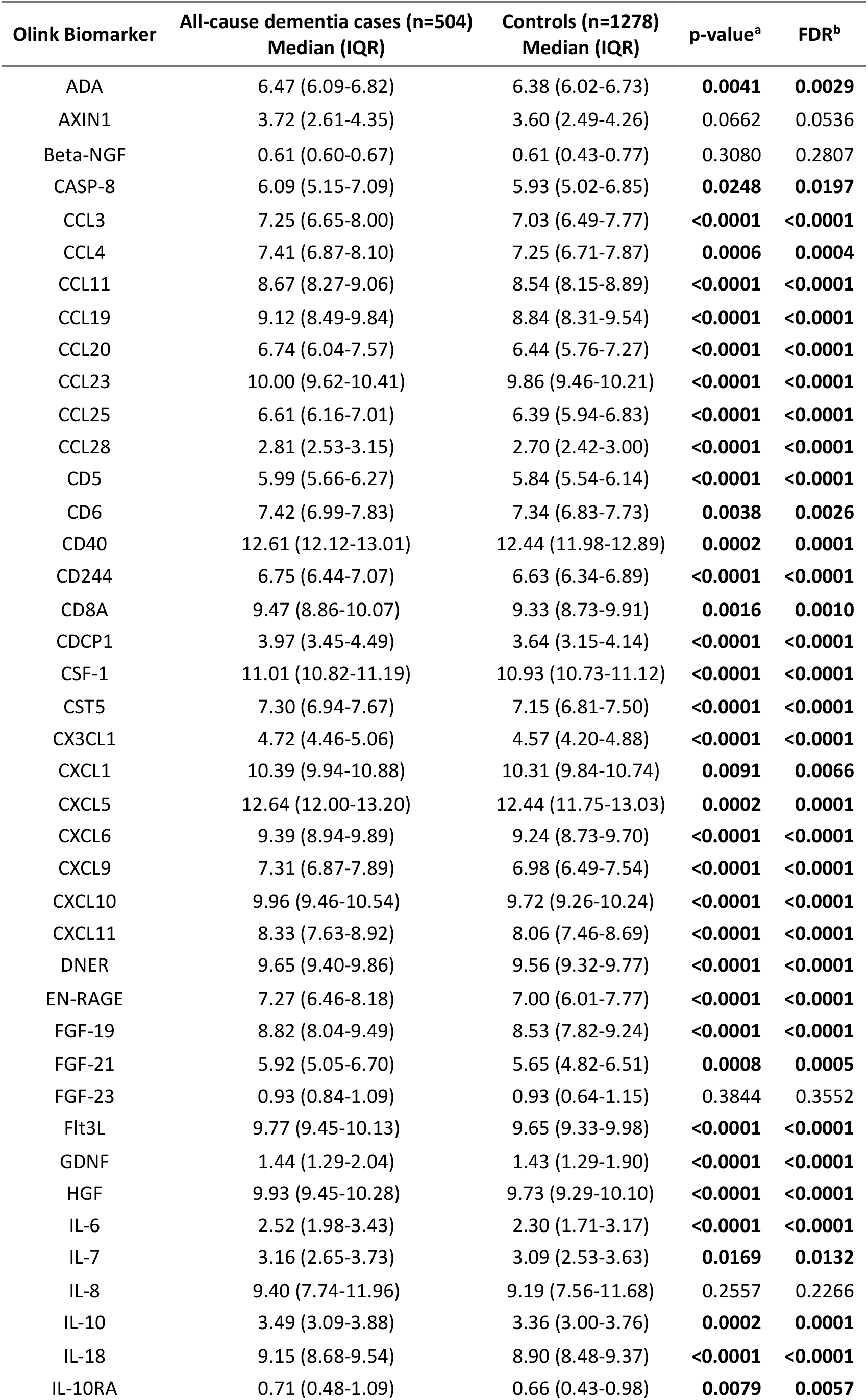
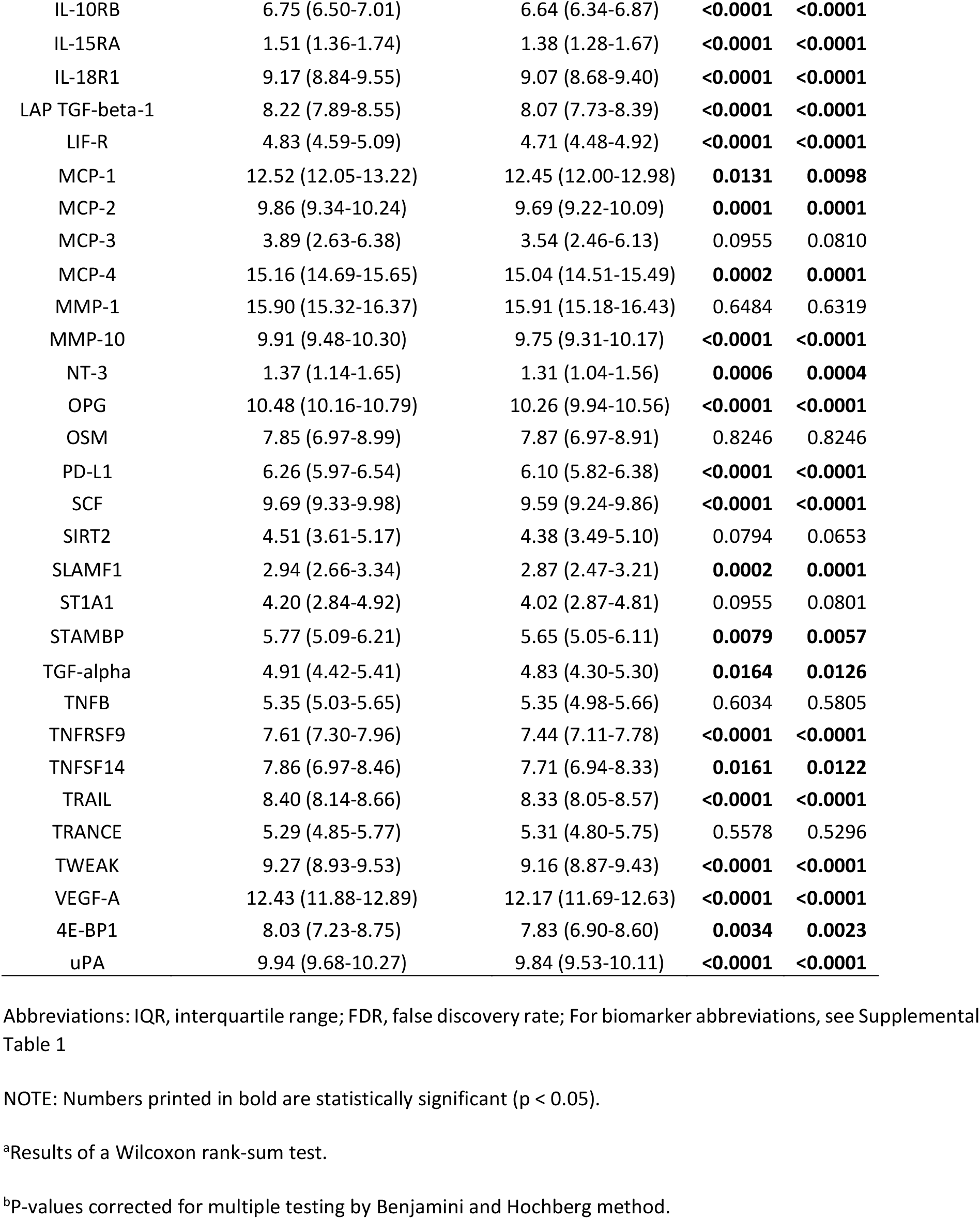
Comparison of median biomarker levels between all-cause dementia cases (n=504) and controls (n=1278)

**Supplemental Table 3.** Comparison of median biomarker levels between Alzheimer’s disease cases (n=163) and controls (n=1278)

| Olink Biomarker | Alzheimer's disease (n=163)<br>Median (IQR) | Controls (n=1278)<br>Median (IQR) | p-value<sup>a</sup> | FDR<sup>b</sup> |
| --- | --- | --- | --- | --- |
| ADA | 6.44 (6.06-6.76) | 6.38 (6.02-6.73) | 0.2526 | 0.3041 |
| AXIN1 | 3.80 (2.89-4.36) | 3.60 (2.49-4.26) | **0.0197** | **0.0331** |
| Beta-NGF | 0.61 (0.56-0.74) | 0.61 (0.43-0.77) | 0.3371 | 0.3916 |
| CASP-8 | 6.29 (5.31-7.44) | 5.93 (5.02-6.85) | **0.0012** | **0.0037** |
| CCL3 | 7.17 (6.60-7.96) | 7.03 (6.49-7.77) | 0.0508 | 0.0704 |
| CCL4 | 7.36 (6.89-8.12) | 7.25 (6.71-7.87) | **0.0363** | 0.0522 |
| CCL11 | 8.67 (8.29-9.01) | 8.54 (8.15-8.89) | **0.0007** | **0.0031** |
| CCL19 | 9.28 (8.60-9.82) | 8.84 (8.31-9.54) | **0.0002** | **0.0010** |
| CCL20 | 6.81 (6.04-7.61) | 6.44 (5.76-7.27) | **0.0018** | **0.0050** |
| CCL23 | 10.03 (9.63-10.41) | 9.86 (9.46-10.21) | **<0.0001** | **0.0004** |
| CCL25 | 6.51 (6.14-6.92) | 6.39 (5.94-6.83) | **0.0099** | **0.0190** |
| CCL28 | 2.83 (2.54-3.24) | 2.70 (2.42-3.00) | **<0.0001** | **0.0004** |
| CD5 | 5.99 (5.70-6.25) | 5.84 (5.54-6.14) | **0.0003** | **0.0013** |
| CD6 | 7.43 (7.05-7.86) | 7.34 (6.83-7.73) | **0.0027** | **0.0069** |
| CD40 | 12.55 (12.18-13.03) | 12.44 (11.98-12.89) | **0.0040** | **0.0093** |
| CD244 | 6.76 (6.45-7.10) | 6.63 (6.34-6.89) | **0.0001** | **0.0009** |
| CD8A | 9.45 (8.80-10.00) | 9.33 (8.73-9.91) | 0.1043 | 0.1350 |
| CDCP1 | 3.94 (3.40-4.40) | 3.64 (3.15-4.14) | **0.0001** | **0.0009** |
| CSF-1 | 11.02 (10.82-11.16) | 10.93 (10.73-11.12) | **0.0029** | **0.0070** |
| CST5 | 7.25 (6.95-7.65) | 7.15 (6.81-7.50) | **0.0009** | **0.0031** |
| CX3CL1 | 4.73 (4.46-5.03) | 4.57 (4.20-4.88) | **<0.0001** | **0.0001** |
| CXCL1 | 10.38 (9.88-10.96) | 10.31 (9.84-10.74) | 0.0872 | 0.1148 |
| CXCL5 | 12.63 (12.09-13.25) | 12.44 (11.75-13.03) | **0.0008** | **0.0031** |
| CXCL6 | 9.39 (8.95-9.91) | 9.24 (8.73-9.70) | **0.0045** | **0.0101** |
| CXCL9 | 7.36 (6.87-7.91) | 6.98 (6.49-7.54) | **<0.0001** | **<0.0001** |
| CXCL10 | 9.90 (9.43-10.61) | 9.72 (9.26-10.24) | **0.0012** | **0.0037** |
| CXCL11 | 8.39 (7.69-8.99) | 8.06 (7.46-8.69) | **0.0008** | **0.0031** |
| DNER | 9.67 (9.40-9.84) | 9.56 (9.32-9.77) | **0.0021** | **0.0057** |
| EN-RAGE | 7.33 (6.56-8.17) | 7.00 (6.01-7.77) | **<0.0001** | **0.0005** |
| FGF-19 | 8.82 (8.09-9.61) | 8.53 (7.82-9.24) | **0.0017** | **0.0050** |
| FGF-21 | 5.93 (5.00-6.70) | 5.65 (4.82-6.51) | **0.0478** | 0.0674 |
| FGF-23 | 0.93 (0.79-1.04) | 0.93 (0.64-1.15) | 0.6932 | 0.7301 |
| Flt3L | 9.76 (9.42-10.11) | 9.65 (9.33-9.98) | **0.0050** | **0.0107** |
| GDNF | 1.44 (1.29-1.99) | 1.43 (1.29-1.90) | **0.0253** | **0.0391** |
| HGF | 9.89 (9.49-10.26) | 9.73 (9.29-10.10) | **0.0006** | **0.0027** |
| IL-6 | 2.52 (1.90-3.60) | 2.30 (1.71-3.17) | **0.0139** | **0.0249** |
| IL-7 | 3.05 (2.48-3.44) | 3.09 (2.53-3.63) | 0.4597 | 0.5114 |
| IL-8 | 9.38 (7.64-12.41) | 9.19 (7.56-11.68) | 0.4592 | 0.5114 |
| IL-10 | 3.48 (3.08-3.84) | 3.36 (3.00-3.76) | **0.0331** | **0.0484** |
| IL-18 | 9.02 (8.64-9.45) | 8.90 (8.48-9.37) | **0.0134** | **0.0247** |
| IL-10RA | 0.73 (0.50-1.07) | 0.66 (0.43-0.98) | **0.0322** | **0.0479** |
| IL-10RB | 6.77 (6.48-6.93) | 6.64 (6.34-6.87) | **0.0001** | **0.0008** |
| IL-15RA | 1.46 (1.36-1.70) | 1.38 (1.28-1.67) | **0.0244** | **0.0385** |
| IL-18R1 | 9.14 (8.80-9.50) | 9.07 (8.68-9.40) | **0.0151** | **0.0265** |
| LAP TGF-beta-1 | 8.30 (7.91-8.61) | 8.07 (7.73-8.39) | **<0.0001** | **0.0001** |
| LIF-R | 4.81 (4.50-5.04) | 4.71 (4.48-4.92) | **0.0008** | **0.0031** |
| MCP-1 | 12.49 (12.04-13.22) | 12.45 (12.00-12.98) | 0.1333 | 0.1699 |
| MCP-2 | 9.82 (9.29-10.15) | 9.69 (9.22-10.09) | 0.0828 | 0.1109 |
| MCP-3 | 3.57 (2.43-5.93) | 3.54 (2.46-6.13) | 0.9037 | 0.9271 |
| MCP-4 | 15.15 (14.71-15.68) | 15.04 (14.51-15.49) | **0.0173** | **0.0297** |
| MMP-1 | 15.89 (15.36-16.38) | 15.91 (15.18-16.43) | 0.5209 | 0.5561 |
| MMP-10 | 9.90 (9.48-10.30) | 9.75 (9.31-10.17) | **0.0029** | **0.0070** |
| NT-3 | 1.32 (1.13-1.59) | 1.31 (1.04-1.56) | 0.4101 | 0.4695 |
| OPG | 10.44 (10.15-10.75) | 10.26 (9.94-10.56) | **<0.0001** | **<0.0001** |
| OSM | 7.98 (6.94-9.06) | 7.87 (6.97-8.91) | 0.5066 | 0.5482 |
| PD-L1 | 6.25 (5.98-6.54) | 6.10 (5.82-6.38) | **<0.0001** | **0.0001** |
| SCF | 9.72 (9.38-9.99) | 9.59 (9.24-9.86) | **0.0002** | **0.0010** |
| SIRT2 | 4.52 (3.79-5.18) | 4.38 (3.49-5.10) | **0.0301** | **0.0458** |
| SLAMF1 | 2.90 (2.51-3.31) | 2.87 (2.47-3.21) | 0.3032 | 0.3575 |
| ST1A1 | 4.37 (3.30-5.04) | 4.02 (2.87-4.81) | **0.0074** | **0.0151** |
| STAMBP | 5.82 (5.22-6.21) | 5.65 (5.05-6.11) | **0.0093** | **0.0184** |
| TGF-alpha | 4.92 (4.48-5.33) | 4.83 (4.30-5.30) | 0.0555 | 0.0756 |
| TNFB | 5.39 (5.08-5.69) | 5.35 (4.98-5.66) | 0.2404 | 0.2967 |
| TNFRSF9 | 7.60 (7.35-7.95) | 7.44 (7.11-7.78) | **<0.0001** | **0.0003** |
| TNFSF14 | 7.90 (7.27-8.49) | 7.71 (6.94-8.33) | **0.0102** | **0.0193** |
| TRAIL | 8.43 (8.13-8.63) | 8.33 (8.05-8.57) | **0.0054** | **0.0113** |
| TRANCE | 5.32 (4.92-5.79) | 5.31 (4.80-5.75) | 0.2541 | 0.3041 |
| TWEAK | 9.30 (8.94-9.58) | 9.16 (8.87-9.43) | **0.0025** | **0.0065** |
| VEGF-A | 12.36 (11.87-12.76) | 12.17 (11.69-12.63) | **0.0012** | **0.0037** |
| 4E-BP1 | 8.07 (7.29-8.68) | 7.83 (6.90-8.60) | **0.0219** | **0.0361** |
| uPA | 9.97 (9.67-10.30) | 9.84 (9.53-10.11) | **<0.0001** | **0.0004** |

Abbreviations: IQR, interquartile range; FDR, false discovery rate; For biomarker abbreviations, see Supplemental Table 1
NOTE: Numbers printed in bold are statistically significant ( $p < 0.05$ ).
<sup>a</sup>Results of a Wilcoxon rank-sum test. Study participants with other (e.g. VD) or unknown dementia forms were excluded.
<sup>b</sup>P-values corrected for multiple testing by Benjamini and Hochberg method.

**Supplemental Table 4.** Comparison of median biomarker levels between vascular dementia cases (n=195) and controls (n=1278)

| Olink Biomarker | Vascular dementia (n=195)<br>Median (IQR) | Controls (n=1278)<br>Median (IQR) | p-value<sup>a</sup> | FDR<sup>b</sup> |
| --- | --- | --- | --- | --- |
| ADA | 6.45 (6.06-6.75) | 6.38 (6.02-6.73) | 0.2154 | 0.2701 |
| AXIN1 | 3.70 (2.39-4.37) | 3.60 (2.49-4.26) | 0.5743 | 0.6312 |
| Beta-NGF | 0.61 (0.59-0.67) | 0.61 (0.43-0.77) | 0.4956 | 0.5758 |
| CASP-8 | 6.04 (5.12-7.10) | 5.93 (5.02-6.85) | 0.2717 | 0.3302 |
| CCL3 | 7.30 (6.76-8.11) | 7.03 (6.49-7.77) | **0.0006** | **0.0015** |
| CCL4 | 7.53 (6.88-8.10) | 7.25 (6.71-7.87) | **0.0025** | **0.0050** |
| CCL11 | 8.68 (8.27-9.06) | 8.54 (8.15-8.89) | **<0.0001** | **0.0003** |
| CCL19 | 9.06 (8.44-9.73) | 8.84 (8.31-9.54) | **0.0114** | **0.0166** |
| CCL20 | 6.79 (6.03-7.74) | 6.44 (5.76-7.27) | **<0.0001** | **0.0002** |
| CCL23 | 9.98 (9.63-10.44) | 9.86 (9.46-10.21) | **<0.0001** | **0.0004** |
| CCL25 | 6.63 (6.09-7.06) | 6.39 (5.94-6.83) | **0.0001** | **0.0004** |
| CCL28 | 2.82 (2.55-3.07) | 2.70 (2.42-3.00) | **0.0005** | **0.0014** |
| CD5 | 6.06 (5.71-6.31) | 5.84 (5.54-6.14) | **<0.0001** | **<0.0001** |
| CD6 | 7.45 (6.97-7.78) | 7.34 (6.83-7.73) | 0.0529 | 0.0733 |
| CD40 | 12.62 (12.12-13.03) | 12.44 (11.98-12.89) | **0.0069** | **0.0106** |
| CD244 | 6.77 (6.44-7.09) | 6.63 (6.34-6.89) | **<0.0001** | **0.0002** |
| CD8A | 9.49 (8.95-10.11) | 9.33 (8.73-9.91) | **0.0057** | **0.0091** |
| CDCP1 | 4.01 (3.47-4.61) | 3.64 (3.15-4.14) | **<0.0001** | **<0.0001** |
| CSF-1 | 11.03 (10.85-11.19) | 10.93 (10.73-11.12) | **<0.0001** | **0.0004** |
| CST5 | 7.28 (6.94-7.64) | 7.15 (6.81-7.50) | **0.0008** | **0.0019** |
| CX3CL1 | 4.71 (4.42-5.05) | 4.57 (4.20-4.88) | **<0.0001** | **<0.0001** |
| CXCL1 | 10.48 (10.07-10.89) | 10.31 (9.84-10.74) | **0.0027** | **0.0051** |
| CXCL5 | 12.69 (12.04-13.21) | 12.44 (11.75-13.03) | **0.0015** | **0.0032** |
| CXCL6 | 9.38 (8.99-9.91) | 9.24 (8.73-9.70) | **0.0006** | **0.0015** |
| CXCL9 | 7.34 (6.89-7.98) | 6.98 (6.49-7.54) | **<0.0001** | **<0.0001** |
| CXCL10 | 10.00 (9.50-10.73) | 9.72 (9.26-10.24) | **<0.0001** | **<0.0001** |
| CXCL11 | 8.30 (7.68-8.89) | 8.06 (7.46-8.69) | **0.0015** | **0.0032** |
| DNER | 9.64 (9.39-9.87) | 9.56 (9.32-9.77) | **0.0019** | **0.0040** |
| EN-RAGE | 7.22 (6.48-8.18) | 7.00 (6.01-7.77) | **0.0002** | **0.0005** |
| FGF-19 | 8.91 (8.02-9.48) | 8.53 (7.82-9.24) | **0.002** | **0.0041** |
| FGF-21 | 5.94 (5.18-6.71) | 5.65 (4.82-6.51) | **0.0028** | **0.0053** |
| FGF-23 | 0.93 (0.79-1.02) | 0.93 (0.64-1.15) | 0.8527 | 0.8749 |
| Flt3L | 9.78 (9.47-10.15) | 9.65 (9.33-9.98) | **0.0002** | **0.0005** |
| GDNF | 1.42 (1.29-2.06) | 1.43 (1.29-1.90) | **0.0063** | **0.0100** |
| HGF | 9.97 (9.50-10.26) | 9.73 (9.29-10.10) | **<0.0001** | **<0.0001** |
| IL-6 | 2.57 (1.99-3.43) | 2.30 (1.71-3.17) | **0.0012** | **0.0026** |
| IL-7 | 3.28 (2.75-3.78) | 3.09 (2.53-3.63) | **0.0031** | **0.0056** |
| IL-8 | 9.42 (7.98-12.28) | 9.19 (7.56-11.68) | 0.1985 | 0.2529 |
| IL-10 | 3.53 (3.11-3.97) | 3.36 (3.00-3.76) | **0.0006** | **0.0015** |
| IL-18 | 9.16 (8.69-9.55) | 8.90 (8.48-9.37) | **<0.0001** | **0.0002** |
| IL-10RA | 0.71 (0.47-1.10) | 0.66 (0.43-0.98) | 0.0681 | 0.0928 |
| IL-10RB | 6.74 (6.48-7.01) | 6.64 (6.34-6.87) | **<0.0001** | **0.0004** |
| IL-15RA | 1.55 (1.36-1.73) | 1.38 (1.28-1.67) | **<0.0001** | **0.0003** |
| IL-18R1 | 9.20 (8.87-9.56) | 9.07 (8.68-9.40) | **<0.0001** | **0.0002** |
| LAP TGF-beta-1 | 8.20 (7.89-8.50) | 8.07 (7.73-8.39) | **0.0001** | **0.0004** |
| LIF-R | 4.83 (4.62-5.10) | 4.71 (4.48-4.92) | **<0.0001** | **<0.0001** |
| MCP-1 | 12.54 (11.99-13.21) | 12.45 (12.00-12.98) | 0.0722 | 0.0967 |
| MCP-2 | 9.92 (9.38-10.30) | 9.69 (9.22-10.09) | **0.0003** | **0.0009** |
| MCP-3 | 3.85 (2.65-6.56) | 3.54 (2.46-6.13) | 0.1077 | 0.1419 |
| MCP-4 | 15.15 (14.70-15.68) | 15.04 (14.51-15.49) | **0.0049** | **0.0082** |
| MMP-1 | 15.91 (15.27-16.36) | 15.91 (15.18-16.43) | 0.9038 | 0.9154 |
| MMP-10 | 9.95 (9.57-10.42) | 9.75 (9.31-10.17) | **<0.0001** | **<0.0001** |
| NT-3 | 1.40 (1.13-1.65) | 1.31 (1.04-1.56) | **0.0044** | **0.0075** |
| OPG | 10.48 (10.15-10.79) | 10.26 (9.94-10.56) | **<0.0001** | **<0.0001** |
| OSM | 7.77 (7.09-9.10) | 7.87 (6.97-8.91) | 0.7958 | 0.8382 |
| PD-L1 | 6.27 (5.95-6.58) | 6.10 (5.82-6.38) | **<0.0001** | **<0.0001** |
| SCF | 9.70 (9.32-9.99) | 9.59 (9.24-9.86) | **0.0050** | **0.0082** |
| SIRT2 | 4.51 (3.45-5.08) | 4.38 (3.49-5.10) | 0.9709 | 0.9709 |
| SLAMF1 | 2.93 (2.66-3.34) | 2.87 (2.47-3.21) | **0.0096** | **0.0143** |
| ST1A1 | 4.13 (2.70-4.85) | 4.02 (2.87-4.81) | 0.5753 | 0.6312 |
| STAMBP | 5.71 (5.04-6.13) | 5.65 (5.05-6.11) | 0.6045 | 0.6541 |
| TGF-alpha | 4.95 (4.49-5.51) | 4.83 (4.30-5.30) | **0.0192** | **0.0275** |
| TNFB | 5.35 (5.01-5.62) | 5.35 (4.98-5.66) | 0.8368 | 0.8698 |
| TNFRSF9 | 7.63 (7.29-8.01) | 7.44 (7.11-7.78) | **<0.0001** | **<0.0001** |
| TNFSF14 | 7.89 (6.87-8.49) | 7.71 (6.94-8.33) | 0.0518 | 0.0731 |
| TRAIL | 8.38 (8.14-8.68) | 8.33 (8.05-8.57) | **0.0030** | **0.0055** |
| TRANCE | 5.29 (4.83-5.86) | 5.31 (4.80-5.75) | 0.3947 | 0.4725 |
| TWEAK | 9.25 (8.92-9.54) | 9.16 (8.87-9.43) | **0.0033** | **0.0058** |
| VEGF-A | 12.45 (11.87-12.95) | 12.17 (11.69-12.63) | **<0.0001** | **<0.0001** |
| 4E-BP1 | 8.00 (7.21-8.65) | 7.83 (6.90-8.60) | 0.2272 | 0.2805 |
| uPA | 9.94 (9.66-10.28) | 9.84 (9.53-10.11) | **0.0003** | **0.0009** |

Abbreviations: IQR, interquartile range; FDR, false discovery rate; For biomarker abbreviations, see Supplemental Table 1
NOTE: Numbers printed in bold are statistically significant ( $p < 0.05$ ).
<sup>a</sup>Results of a Wilcoxon rank-sum test. Study participants with other (e.g. AD) or unknown dementia forms were excluded.
<sup>b</sup>P-values corrected for multiple testing by Benjamini and Hochberg method.

**Supplemental Table 5.** Non-significant associations of Olink Inflammation panel biomarker levels with **all-cause dementia** incidence (FDR ≥ 0.05)

| Olink Biomarker | Value of 1 SD | All-cause dementia (n=504 cases) |  |  |
| --- | --- | --- | --- | --- |
|  |  | OR (95% CI) per 1 SD<sup>a</sup> | p-value per 1 SD | FDR corrected p-value<sup>b</sup> |
| Beta-NGF | 0.397 | 0.97 (0.86-1.11) | 0.6887 | 0.6984 |
| CD8A | 0.944 | 1.13 (1.00-1.27) | **0.0492** | 0.0581 |
| FGF-21 | 1.315 | 1.11 (0.99-1.25) | 0.0716 | 0.0818 |
| FGF-23 | 0.449 | 0.99 (0.88-1.11) | 0.8587 | 0.8587 |
| IL-6 | 1.871 | 1.10 (0.98-1.23) | 0.1092 | 0.1210 |
| IL-8 | 2.570 | 1.09 (0.97-1.22) | 0.1471 | 0.1605 |
| IL-12B | 0.795 | 1.10 (0.98-1.24) | 0.1086 | 0.1210 |
| MCP-1 | 1.001 | 1.12 (1.00-1.26) | **0.0469** | 0.0563 |
| MCP-3 | 2.846 | 1.05 (0.94-1.18) | 0.3626 | 0.3784 |
| MMP-1 | 0.981 | 1.04 (0.93-1.18) | 0.4651 | 0.4784 |
| OSM | 1.501 | 1.08 (0.96-1.21) | 0.2165 | 0.2292 |
| SLAMF1 | 0.633 | 1.12 (1.00-1.27) | 0.0506 | 0.0588 |
| TNFB | 0.592 | 1.08 (0.96-1.22) | 0.1823 | 0.1959 |
| 4E-BP1 | 1.300 | 1.13 (1.00-1.27) | **0.0457** | 0.0558 |

Abbreviations: SD, standard deviation; CI, confidence interval; FDR, false discovery rate; For biomarker abbreviations, see Supplemental Table 1.
<sup>a</sup> Multivariate logistic regression model adjusted for age (continuously), sex, education, physical activity, BMI (categorical), CVD, diabetes, depression, *APOE* genotype.
<sup>b</sup> P-values corrected for multiple testing by the Benjamini and Hochberg method.

**Supplemental Table 6.**
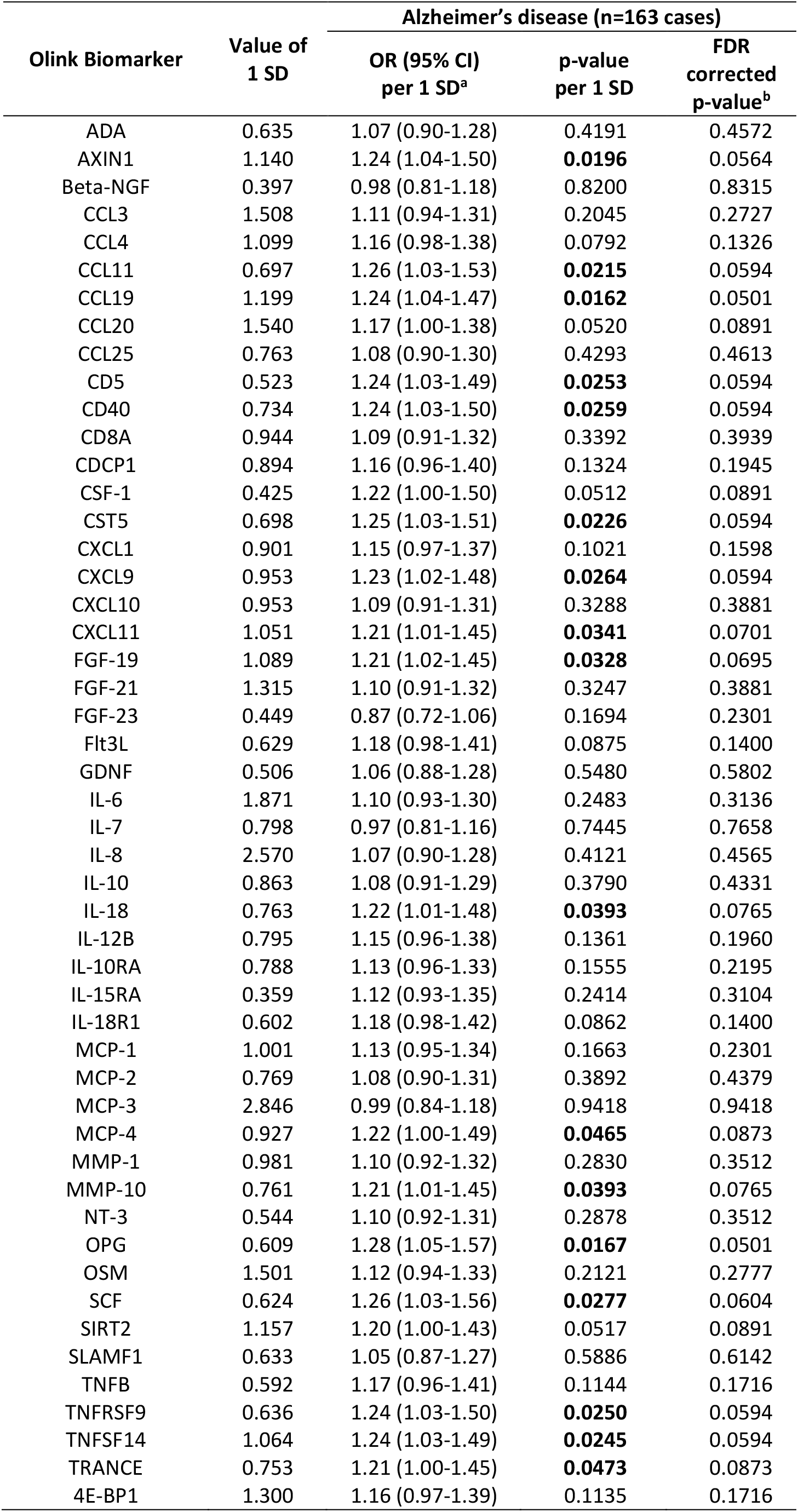
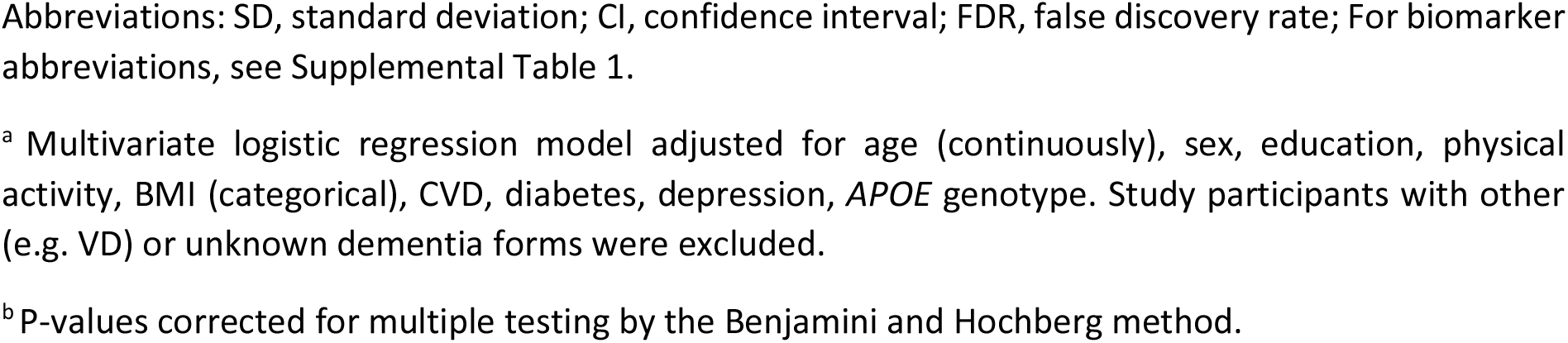
Non-significant associations of Olink Inflammation panel biomarker levels with **Alzheimer’s disease** incidence (FDR ≥ 0.05)

| Olink Biomarker | Value of 1 SD | Alzheimer's disease (n=163 cases) |  |  |
| --- | --- | --- | --- | --- |
|  |  | OR (95% CI) per 1 SD<sup>a</sup> | p-value per 1 SD | FDR corrected p-value<sup>b</sup> |
| ADA | 0.635 | 1.07 (0.90-1.28) | 0.4191 | 0.4572 |
| AXIN1 | 1.140 | 1.24 (1.04-1.50) | **0.0196** | 0.0564 |
| Beta-NGF | 0.397 | 0.98 (0.81-1.18) | 0.8200 | 0.8315 |
| CCL3 | 1.508 | 1.11 (0.94-1.31) | 0.2045 | 0.2727 |
| CCL4 | 1.099 | 1.16 (0.98-1.38) | 0.0792 | 0.1326 |
| CCL11 | 0.697 | 1.26 (1.03-1.53) | **0.0215** | 0.0594 |
| CCL19 | 1.199 | 1.24 (1.04-1.47) | **0.0162** | 0.0501 |
| CCL20 | 1.540 | 1.17 (1.00-1.38) | 0.0520 | 0.0891 |
| CCL25 | 0.763 | 1.08 (0.90-1.30) | 0.4293 | 0.4613 |
| CD5 | 0.523 | 1.24 (1.03-1.49) | **0.0253** | 0.0594 |
| CD40 | 0.734 | 1.24 (1.03-1.50) | **0.0259** | 0.0594 |
| CD8A | 0.944 | 1.09 (0.91-1.32) | 0.3392 | 0.3939 |
| CDCP1 | 0.894 | 1.16 (0.96-1.40) | 0.1324 | 0.1945 |
| CSF-1 | 0.425 | 1.22 (1.00-1.50) | 0.0512 | 0.0891 |
| CST5 | 0.698 | 1.25 (1.03-1.51) | **0.0226** | 0.0594 |
| CXCL1 | 0.901 | 1.15 (0.97-1.37) | 0.1021 | 0.1598 |
| CXCL9 | 0.953 | 1.23 (1.02-1.48) | **0.0264** | 0.0594 |
| CXCL10 | 0.953 | 1.09 (0.91-1.31) | 0.3288 | 0.3881 |
| CXCL11 | 1.051 | 1.21 (1.01-1.45) | **0.0341** | 0.0701 |
| FGF-19 | 1.089 | 1.21 (1.02-1.45) | **0.0328** | 0.0695 |
| FGF-21 | 1.315 | 1.10 (0.91-1.32) | 0.3247 | 0.3881 |
| FGF-23 | 0.449 | 0.87 (0.72-1.06) | 0.1694 | 0.2301 |
| Flt3L | 0.629 | 1.18 (0.98-1.41) | 0.0875 | 0.1400 |
| GDNF | 0.506 | 1.06 (0.88-1.28) | 0.5480 | 0.5802 |
| IL-6 | 1.871 | 1.10 (0.93-1.30) | 0.2483 | 0.3136 |
| IL-7 | 0.798 | 0.97 (0.81-1.16) | 0.7445 | 0.7658 |
| IL-8 | 2.570 | 1.07 (0.90-1.28) | 0.4121 | 0.4565 |
| IL-10 | 0.863 | 1.08 (0.91-1.29) | 0.3790 | 0.4331 |
| IL-18 | 0.763 | 1.22 (1.01-1.48) | **0.0393** | 0.0765 |
| IL-12B | 0.795 | 1.15 (0.96-1.38) | 0.1361 | 0.1960 |
| IL-10RA | 0.788 | 1.13 (0.96-1.33) | 0.1555 | 0.2195 |
| IL-15RA | 0.359 | 1.12 (0.93-1.35) | 0.2414 | 0.3104 |
| IL-18R1 | 0.602 | 1.18 (0.98-1.42) | 0.0862 | 0.1400 |
| MCP-1 | 1.001 | 1.13 (0.95-1.34) | 0.1663 | 0.2301 |
| MCP-2 | 0.769 | 1.08 (0.90-1.31) | 0.3892 | 0.4379 |
| MCP-3 | 2.846 | 0.99 (0.84-1.18) | 0.9418 | 0.9418 |
| MCP-4 | 0.927 | 1.22 (1.00-1.49) | **0.0465** | 0.0873 |
| MMP-1 | 0.981 | 1.10 (0.92-1.32) | 0.2830 | 0.3512 |
| MMP-10 | 0.761 | 1.21 (1.01-1.45) | **0.0393** | 0.0765 |
| NT-3 | 0.544 | 1.10 (0.92-1.31) | 0.2878 | 0.3512 |
| OPG | 0.609 | 1.28 (1.05-1.57) | **0.0167** | 0.0501 |
| OSM | 1.501 | 1.12 (0.94-1.33) | 0.2121 | 0.2777 |
| SCF | 0.624 | 1.26 (1.03-1.56) | **0.0277** | 0.0604 |
| SIRT2 | 1.157 | 1.20 (1.00-1.43) | 0.0517 | 0.0891 |
| SLAMF1 | 0.633 | 1.05 (0.87-1.27) | 0.5886 | 0.6142 |
| TNFB | 0.592 | 1.17 (0.96-1.41) | 0.1144 | 0.1716 |
| TNFRSF9 | 0.636 | 1.24 (1.03-1.50) | **0.0250** | 0.0594 |
| TNFSF14 | 1.064 | 1.24 (1.03-1.49) | **0.0245** | 0.0594 |
| TRANCE | 0.753 | 1.21 (1.00-1.45) | **0.0473** | 0.0873 |
| 4E-BP1 | 1.300 | 1.16 (0.97-1.39) | 0.1135 | 0.1716 |

Abbreviations: SD, standard deviation; CI, confidence interval; FDR, false discovery rate; For biomarker abbreviations, see Supplemental Table 1.
<sup>a</sup> Multivariate logistic regression model adjusted for age (continuously), sex, education, physical activity, BMI (categorical), CVD, diabetes, depression, *APOE* genotype. Study participants with other (e.g. VD) or unknown dementia forms were excluded.
<sup>b</sup> P-values corrected for multiple testing by the Benjamini and Hochberg method.

**Supplemental Table 7.**
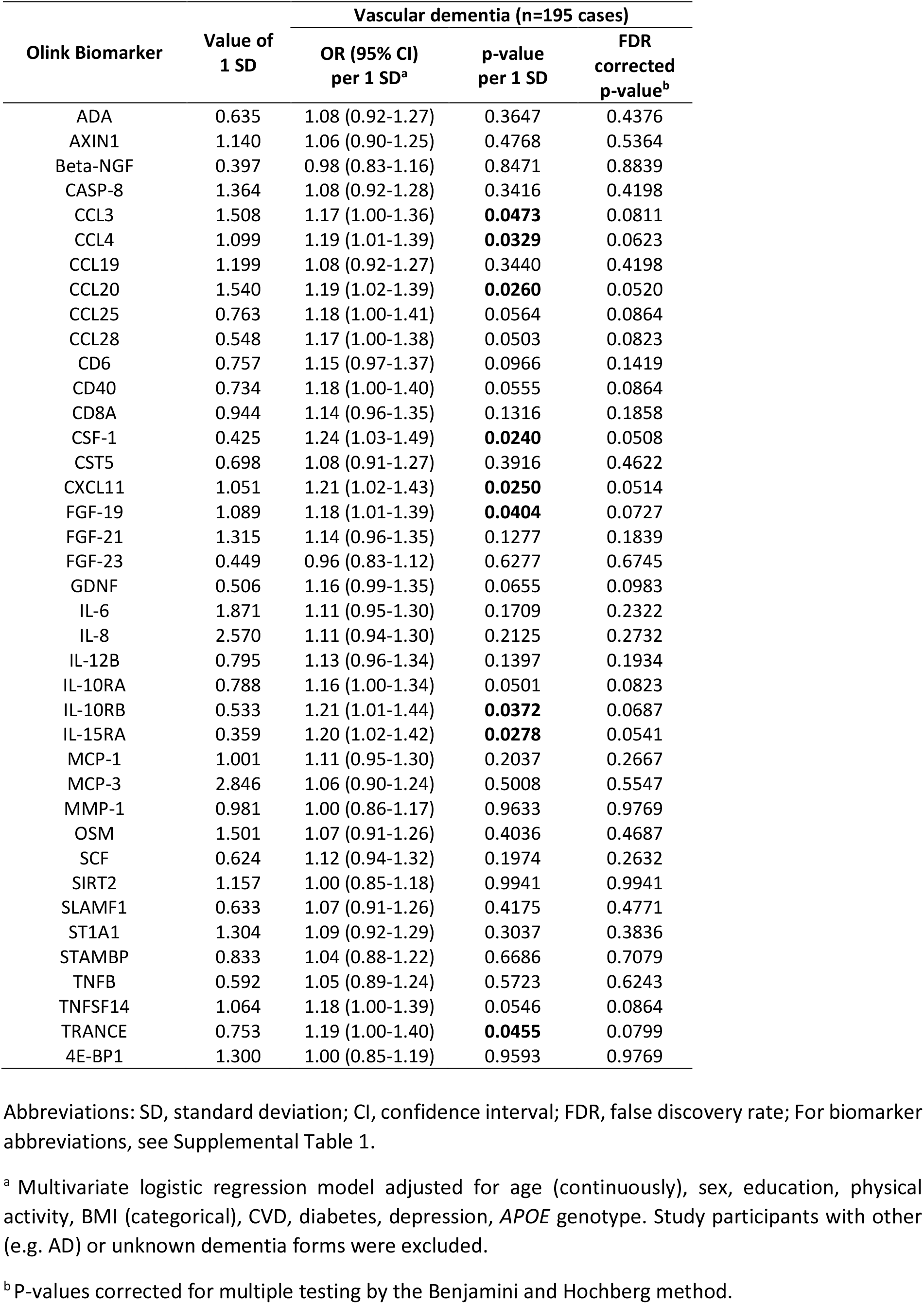
Non-significant associations of Olink Inflammation panel biomarker levels with **vascular dementia** incidence (FDR ≥ 0.05)

| Olink Biomarker | Value of 1 SD | Vascular dementia (n=195 cases) |  |  |
| --- | --- | --- | --- | --- |
|  |  | OR (95% CI) per 1 SD<sup>a</sup> | p-value per 1 SD | FDR corrected p-value<sup>b</sup> |
| ADA | 0.635 | 1.08 (0.92-1.27) | 0.3647 | 0.4376 |
| AXIN1 | 1.140 | 1.06 (0.90-1.25) | 0.4768 | 0.5364 |
| Beta-NGF | 0.397 | 0.98 (0.83-1.16) | 0.8471 | 0.8839 |
| CASP-8 | 1.364 | 1.08 (0.92-1.28) | 0.3416 | 0.4198 |
| CCL3 | 1.508 | 1.17 (1.00-1.36) | **0.0473** | 0.0811 |
| CCL4 | 1.099 | 1.19 (1.01-1.39) | **0.0329** | 0.0623 |
| CCL19 | 1.199 | 1.08 (0.92-1.27) | 0.3440 | 0.4198 |
| CCL20 | 1.540 | 1.19 (1.02-1.39) | **0.0260** | 0.0520 |
| CCL25 | 0.763 | 1.18 (1.00-1.41) | 0.0564 | 0.0864 |
| CCL28 | 0.548 | 1.17 (1.00-1.38) | 0.0503 | 0.0823 |
| CD6 | 0.757 | 1.15 (0.97-1.37) | 0.0966 | 0.1419 |
| CD40 | 0.734 | 1.18 (1.00-1.40) | 0.0555 | 0.0864 |
| CD8A | 0.944 | 1.14 (0.96-1.35) | 0.1316 | 0.1858 |
| CSF-1 | 0.425 | 1.24 (1.03-1.49) | **0.0240** | 0.0508 |
| CST5 | 0.698 | 1.08 (0.91-1.27) | 0.3916 | 0.4622 |
| CXCL11 | 1.051 | 1.21 (1.02-1.43) | **0.0250** | 0.0514 |
| FGF-19 | 1.089 | 1.18 (1.01-1.39) | **0.0404** | 0.0727 |
| FGF-21 | 1.315 | 1.14 (0.96-1.35) | 0.1277 | 0.1839 |
| FGF-23 | 0.449 | 0.96 (0.83-1.12) | 0.6277 | 0.6745 |
| GDNF | 0.506 | 1.16 (0.99-1.35) | 0.0655 | 0.0983 |
| IL-6 | 1.871 | 1.11 (0.95-1.30) | 0.1709 | 0.2322 |
| IL-8 | 2.570 | 1.11 (0.94-1.30) | 0.2125 | 0.2732 |
| IL-12B | 0.795 | 1.13 (0.96-1.34) | 0.1397 | 0.1934 |
| IL-10RA | 0.788 | 1.16 (1.00-1.34) | 0.0501 | 0.0823 |
| IL-10RB | 0.533 | 1.21 (1.01-1.44) | **0.0372** | 0.0687 |
| IL-15RA | 0.359 | 1.20 (1.02-1.42) | **0.0278** | 0.0541 |
| MCP-1 | 1.001 | 1.11 (0.95-1.30) | 0.2037 | 0.2667 |
| MCP-3 | 2.846 | 1.06 (0.90-1.24) | 0.5008 | 0.5547 |
| MMP-1 | 0.981 | 1.00 (0.86-1.17) | 0.9633 | 0.9769 |
| OSM | 1.501 | 1.07 (0.91-1.26) | 0.4036 | 0.4687 |
| SCF | 0.624 | 1.12 (0.94-1.32) | 0.1974 | 0.2632 |
| SIRT2 | 1.157 | 1.00 (0.85-1.18) | 0.9941 | 0.9941 |
| SLAMF1 | 0.633 | 1.07 (0.91-1.26) | 0.4175 | 0.4771 |
| ST1A1 | 1.304 | 1.09 (0.92-1.29) | 0.3037 | 0.3836 |
| STAMBP | 0.833 | 1.04 (0.88-1.22) | 0.6686 | 0.7079 |
| TNFB | 0.592 | 1.05 (0.89-1.24) | 0.5723 | 0.6243 |
| TNFSF14 | 1.064 | 1.18 (1.00-1.39) | 0.0546 | 0.0864 |
| TRANCE | 0.753 | 1.19 (1.00-1.40) | **0.0455** | 0.0799 |
| 4E-BP1 | 1.300 | 1.00 (0.85-1.19) | 0.9593 | 0.9769 |

Abbreviations: SD, standard deviation; CI, confidence interval; FDR, false discovery rate; For biomarker abbreviations, see Supplemental Table 1.
<sup>a</sup> Multivariate logistic regression model adjusted for age (continuously), sex, education, physical activity, BMI (categorical), CVD, diabetes, depression, *APOE* genotype. Study participants with other (e.g. AD) or unknown dementia forms were excluded.
<sup>b</sup> P-values corrected for multiple testing by the Benjamini and Hochberg method.

**Supplemental Table 8.**
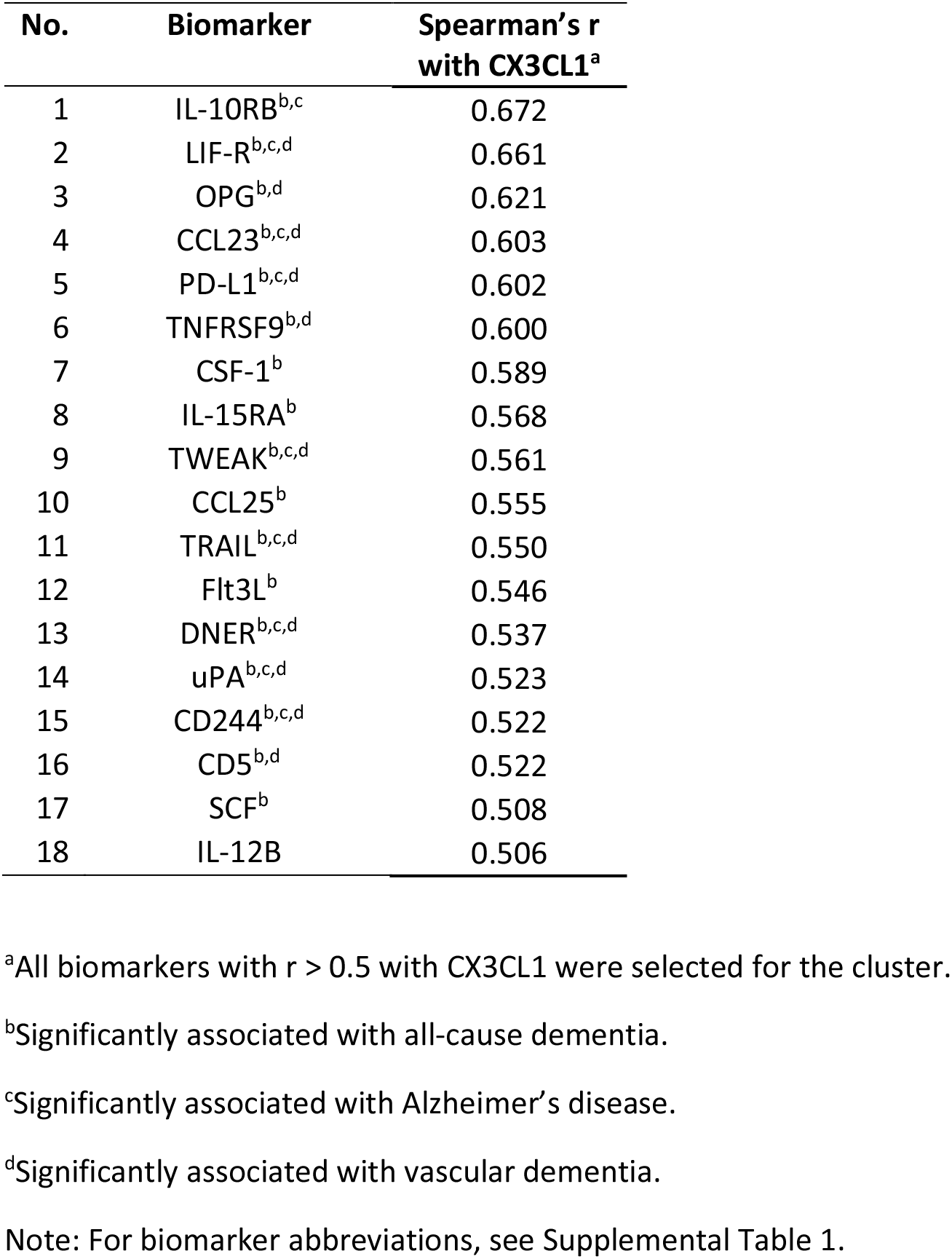
Olink Inflammation Panel Biomarkers in the CX3CL1 cluster

**Supplemental Table 9.**
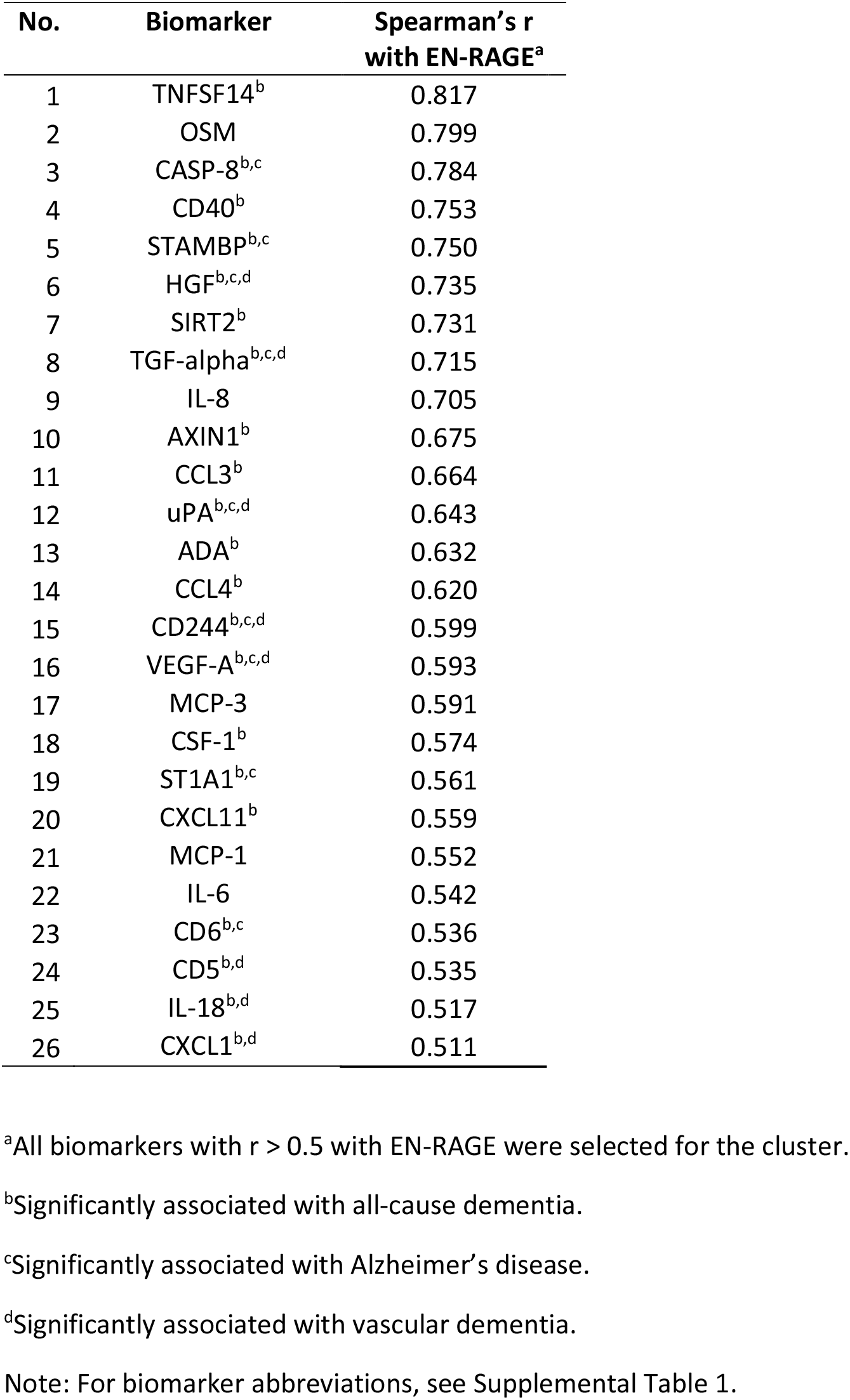
Olink Inflammation Panel Biomarkers in the EN-RAGE cluster

| No. | Biomarker | Spearman's r<br>with EN-RAGE<sup>a</sup> |
| --- | --- | --- |
| 1 | TNFSF14<sup>b</sup> | 0.817 |
| 2 | OSM | 0.799 |
| 3 | CASP-8<sup>b,c</sup> | 0.784 |
| 4 | CD40<sup>b</sup> | 0.753 |
| 5 | STAMBP<sup>b,c</sup> | 0.750 |
| 6 | HGF<sup>b,c,d</sup> | 0.735 |
| 7 | SIRT2<sup>b</sup> | 0.731 |
| 8 | TGF-alpha<sup>b,c,d</sup> | 0.715 |
| 9 | IL-8 | 0.705 |
| 10 | AXIN1<sup>b</sup> | 0.675 |
| 11 | CCL3<sup>b</sup> | 0.664 |
| 12 | uPA<sup>b,c,d</sup> | 0.643 |
| 13 | ADA<sup>b</sup> | 0.632 |
| 14 | CCL4<sup>b</sup> | 0.620 |
| 15 | CD244<sup>b,c,d</sup> | 0.599 |
| 16 | VEGF-A<sup>b,c,d</sup> | 0.593 |
| 17 | MCP-3 | 0.591 |
| 18 | CSF-1<sup>b</sup> | 0.574 |
| 19 | ST1A1<sup>b,c</sup> | 0.561 |
| 20 | CXCL11<sup>b</sup> | 0.559 |
| 21 | MCP-1 | 0.552 |
| 22 | IL-6 | 0.542 |
| 23 | CD6<sup>b,c</sup> | 0.536 |
| 24 | CD5<sup>b,d</sup> | 0.535 |
| 25 | IL-18<sup>b,d</sup> | 0.517 |
| 26 | CXCL1<sup>b,d</sup> | 0.511 |

<sup>a</sup>All biomarkers with $r > 0.5$ with EN-RAGE were selected for the cluster.
<sup>b</sup>Significantly associated with all-cause dementia.
<sup>c</sup>Significantly associated with Alzheimer's disease.
<sup>d</sup>Significantly associated with vascular dementia.
Note: For biomarker abbreviations, see Supplemental Table 1.

**Supplemental Table 10.** Olink Inflammation Panel Biomarkers in the LAP TGF-beta-1 cluster

| No. | Biomarker | Spearman's r with<br>LAP TGF-beta-1<sup>a</sup> |
| --- | --- | --- |
| 1 | HGF<sup>b,c,d</sup> | 0.601 |
| 2 | CD244<sup>b,c,d</sup> | 0.577 |
| 3 | TWEAK<sup>b,c,d</sup> | 0.560 |
| 4 | uPA<sup>b,c,d</sup> | 0.557 |
| 5 | OPG<sup>b,d</sup> | 0.543 |
| 6 | LIF-R<sup>b,c,d</sup> | 0.540 |
| 7 | CSF-1 | 0.539 |
| 8 | IL-10RB<sup>b,c</sup> | 0.539 |
| 9 | PD-L1<sup>b,c,d</sup> | 0.518 |
| 10 | VEGF-A<sup>b,d</sup> | 0.516 |
| 11 | DNER<sup>b,c,d</sup> | 0.513 |
| 12 | CD5<sup>b,d</sup> | 0.510 |
| 13 | CCL11<sup>b,d</sup> | 0.509 |
| 14 | MCP-4<sup>b,d</sup> | 0.504 |
| 15 | CXCL6<sup>b,c,d</sup> | 0.501 |
| 16 | CD40<sup>b</sup> | 0.500 |

<sup>a</sup>All biomarkers with $r > 0.5$ with LAP TGF-beta-1 were selected for the cluster.
<sup>b</sup>Significantly associated with all-cause dementia.
<sup>c</sup>Significantly associated with Alzheimer's disease.
<sup>d</sup>Significantly associated with vascular dementia.
Note: For biomarker abbreviations, see Supplemental Table 1.

**Supplemental Table 11.** Olink Inflammation Panel Biomarkers in the VEGF-A cluster

| No. | Biomarker | Spearman's r<br>with VEGF-A<sup>a</sup> |
| --- | --- | --- |
| 1 | HGF<sup>b,c,d</sup> | 0.680 |
| 2 | CD40<sup>b</sup> | 0.646 |
| 3 | CSF-1 | 0.620 |
| 4 | uPA<sup>b,c,d</sup> | 0.608 |
| 5 | EN-RAGE<sup>b,c,d</sup> | 0.593 |
| 6 | CD244<sup>b,c,d</sup> | 0.584 |
| 7 | MCP-1 | 0.563 |
| 8 | OSM | 0.561 |
| 9 | CXCL1<sup>b,d</sup> | 0.558 |
| 10 | CCL3<sup>b</sup> | 0.556 |
| 11 | TGF-α<sup>b,c,d</sup> | 0.554 |
| 12 | TNFSF14<sup>b</sup> | 0.551 |
| 13 | IL-6 | 0.538 |
| 14 | CCL4<sup>b</sup> | 0.534 |
| 15 | STAMBP<sup>b,c</sup> | 0.531 |
| 16 | PD-L1<sup>b,c,d</sup> | 0.529 |
| 17 | IL-10RB<sup>b,c</sup> | 0.526 |
| 18 | CD5<sup>b,d</sup> | 0.522 |
| 19 | TNFRSF9<sup>b,d</sup> | 0.520 |
| 20 | CXCL11<sup>b</sup> | 0.519 |
| 21 | IL-10<sup>b,d</sup> | 0.518 |
| 22 | OPG<sup>b,d</sup> | 0.518 |
| 23 | SIRT2<sup>b</sup> | 0.518 |
| 24 | LAP TGF-β-1<sup>b,c,d</sup> | 0.516 |
| 25 | ADA<sup>b</sup> | 0.514 |
| 26 | TWEAK<sup>b,c,d</sup> | 0.504 |
| 27 | IL-18R1<sup>b,d</sup> | 0.504 |
| 28 | CCL11<sup>b,d</sup> | 0.502 |

<sup>a</sup>All biomarkers with $r > 0.5$ with VEGF-A were selected for the cluster.
<sup>b</sup>Significantly associated with all-cause dementia.
<sup>c</sup>Significantly associated with Alzheimer's disease.
<sup>d</sup>Significantly associated with vascular dementia.
For biomarker abbreviations, see Supplemental Table 1.

**Supplemental Table 12.** Exploratory subgroup and sensitivity analyses for the association of CX3CL1 and all-cause dementia

| Group | n<sub>total</sub> | n<sub>cases</sub> | OR (95% CI)<sup>a</sup> | p-value interaction |
| --- | --- | --- | --- | --- |
| **Total cohort** | 1782 | 504 | 1.41 (1.24-1.60) |  |
| **Stratified by age** |  |  |  | 0.2767 |
| <68 years | 1258 | 254 | 1.51 (1.29-1.78) |  |
| ≥68 years | 524 | 250 | 1.49 (1.22-1.83) |  |
| **Stratified by sex** |  |  |  | 0.6705 |
| Women | 965 | 262 | 1.36 (1.14-1.61) |  |
| Men | 817 | 242 | 1.48 (1.23-1.78) |  |
| **Stratified by obesity** |  |  |  | 0.9649 |
| BMI < 30 kg/m<sup>2</sup> | 479 | 144 | 1.46 (1.14-1.87) |  |
| BMI ≥ 30 kg/m<sup>2</sup> | 1303 | 360 | 1.40 (1.21-1.62) |  |
| **Stratified by diabetes** |  |  |  | 0.0804 |
| No | 1489 | 394 | 1.47 (1.28-1.69) |  |
| Yes | 293 | 110 | 1.19 (0.87-1.64) |  |
| **Stratified by CVD** |  |  |  | 0.8759 |
| No | 1373 | 350 | 1.40 (1.21-1.63) |  |
| Yes | 409 | 154 | 1.45 (1.13-1.86) |  |
| **Stratified by APOE ε4** |  |  |  | 0.5879 |
| Negative | 1276 | 313 | 1.42 (1.22-1.66) |  |
| Positive | 506 | 193 | 1.34 (1.07-1.69) |  |
| **Stratified by time of diagnosis** |  |  |  | NA |
| In first 10 years of FUP | 1498 | 220 | 1.65 (1.36-2.00) |  |
| In year 11-19 of FUP | 1562 | 284 | 1.28 (1.10-1.48) |  |
| **Excluding subjects free of dementia who died prior to 80<sup>th</sup> birthday** |  |  |  | NA |
| No | 1782 | 504 | 1.41 (1.24-1.60) |  |
| Yes | 1601 | 504 | 1.47 (1.29-1.67) |  |
| **Excluding subjects with sign of acute infection (CRP level >20mg/L)** |  |  |  | NA |
| No | 1782 | 504 | 1.41 (1.24-1.60) |  |
| Yes | 1745 | 497 | 1.41 (1.24-1.60) |  |

Abbreviations: CX3CL1, Fractalkine; OR, odds ratio; CI, confidence interval; CVD, cardiovascular disease; APOE, apolipoprotein E; CRP, C-reactive protein;
NOTE: Numbers printed in bold are statistically significant (p < 0.05).
<sup>a</sup>Results of multivariate logistic regression model adjusted for age (continuously), sex, education, physical activity, BMI (categorical), CVD, diabetes, depression, APOE genotype.

**Supplemental Table 13.** Exploratory subgroup and sensitivity analyses for the association of EN-RAGE and all-cause dementia

| Group | n<sub>total</sub> | n<sub>cases</sub> | OR (95% CI)<sup>a</sup> | p-value interaction |
| --- | --- | --- | --- | --- |
| **Total cohort** | 1782 | 504 | 1.41 (1.25-1.60) |  |
| **Stratified by age** |  |  |  | 0.3201 |
| <68 years | 1258 | 254 | 1.41 (1.21-1.64) |  |
| ≥68 years | 524 | 250 | 1.34 (1.11-1.61) |  |
| **Stratified by sex** |  |  |  | 0.2769 |
| Women | 965 | 262 | 1.55 (1.30-1.85) |  |
| Men | 817 | 242 | 1.32 (1.11-1.56) |  |
| **Stratified by obesity** |  |  |  | 0.2911 |
| BMI < 30 kg/m<sup>2</sup> | 479 | 144 | 1.31 (1.04-1.66) |  |
| BMI ≥ 30 kg/m<sup>2</sup> | 1303 | 360 | 1.48 (1.28-1.71) |  |
| **Stratified by diabetes** |  |  |  | 0.3445 |
| No | 1489 | 394 | 1.45 (1.27-1.66) |  |
| Yes | 293 | 110 | 1.31 (0.99-1.74) |  |
| **Stratified by CVD** |  |  |  | 0.9128 |
| No | 1373 | 350 | 1.43 (1.24-1.66) |  |
| Yes | 409 | 154 | 1.37 (1.09-1.73) |  |
| **Stratified by APOE ε4** |  |  |  | **0.0241** |
| Negative | 1276 | 313 | 1.56 (1.34-1.81) |  |
| Positive | 506 | 193 | 1.16 (0.94-1.42) |  |
| **Stratified by time of diagnosis** |  |  |  | NA |
| In first 10 years of FUP | 1498 | 220 | 1.46 (1.22-1.73) |  |
| In year 11-19 of FUP | 1562 | 284 | 1.36 (1.18-1.57) |  |
| **Excluding subjects free of dementia who died prior to 80<sup>th</sup> birthday** |  |  |  | NA |
| No | 1782 | 504 | 1.41 (1.25-1.60) |  |
| Yes | 1601 | 504 | 1.49 (1.31-1.68) |  |
| **Excluding subjects with sign of acute infection (CRP level >20mg/L)** |  |  |  | NA |
| No | 1782 | 504 | 1.41 (1.25-1.60) |  |
| Yes | 1745 | 497 | 1.43 (1.27-1.62) |  |

Abbreviations: EN-RAGE, Protein S100-A12; OR, odds ratio; CI, confidence interval; CVD, cardiovascular disease; APOE, apolipoprotein E; CRP, C-reactive protein;
NOTE: Numbers printed in bold are statistically significant (p < 0.05).
<sup>a</sup> Results of multivariate logistic regression model adjusted for age (continuously), sex, education, physical activity, BMI (categorical), CVD, diabetes, depression, *APOE* genotype.

**Supplemental Table 14.** Exploratory subgroup and sensitivity analyses for the association of EN-RAGE and Alzheimer’s disease

| Group | n<sub>total</sub> | n<sub>cases</sub> | OR (95% CI)<sup>a</sup> | p-value interaction |
| --- | --- | --- | --- | --- |
| **Total cohort** | 1782 | 163 | 1.51 (1.25-1.83) |  |
| **Stratified by age** |  |  |  | 0.7589 |
| <68 years | 1258 | 88 | 1.43 (1.12-1.81) |  |
| ≥68 years | 524 | 75 | 1.54 (1.12-2.11) |  |
| **Stratified by sex** |  |  |  | 0.6325 |
| Women | 965 | 95 | 1.46 (1.14-1.88) |  |
| Men | 817 | 68 | 1.58 (1.16-2.15) |  |
| **Stratified by obesity** |  |  |  | 0.9321 |
| BMI < 30 kg/m<sup>2</sup> | 479 | 53 | 1.53 (1.06-2.22) |  |
| BMI ≥ 30 kg/m<sup>2</sup> | 1303 | 110 | 1.50 (1.19-1.89) |  |
| **Stratified by diabetes** |  |  |  | 0.2142 |
| No | 1489 | 131 | 1.40 (1.14-1.72) |  |
| Yes | 293 | 32 | 2.01 (1.22-3.30)<sup>b</sup> |  |
| **Stratified by CVD** |  |  |  | 0.4966 |
| No | 1373 | 124 | 1.56 (1.25-1.95) |  |
| Yes | 409 | 39 | 1.31 (0.90-1.91)<sup>b</sup> |  |
| **Stratified by APOE ε4** |  |  |  | 0.0798 |
| Negative | 1276 | 87 | 1.78 (1.35-2.33) |  |
| Positive | 506 | 76 | 1.22 (0.93-1.61) |  |
| **Stratified by time of diagnosis** |  |  |  | NA |
| In first 10 years of FUP | 1699 | 80 | 1.79 (1.34-2.36) |  |
| In year 11-19 of FUP | 1702 | 83 | 1.31 (1.02-1.67) |  |
| **Excluding subjects free of dementia who died prior to 80<sup>th</sup> birthday** |  |  |  | NA |
| No | 1782 | 163 | 1.51 (1.25-1.83) |  |
| Yes | 1601 | 163 | 1.58 (1.30-1.92) |  |
| **Excluding subjects with sign of acute infection (CRP level >20mg/L)** |  |  |  | NA |
| No | 1782 | 163 | 1.51 (1.25-1.83) |  |
| Yes | 1745 | 160 | 1.51 (1.24-1.83) |  |

Abbreviations: EN-RAGE, Protein S100-A12; OR, odds ratio; CI, confidence interval; CVD, cardiovascular disease; APOE, apolipoprotein E; CRP, C-reactive protein;
NOTE: Numbers printed in bold are statistically significant (p < 0.05).
<sup>a</sup>Results of multivariate logistic regression model adjusted for age (continuously), sex, education, physical activity, BMI (categorical), CVD, diabetes, depression, *APOE* genotype. Study participants with other (e.g. VD) or unknown dementia forms were excluded.
<sup>b</sup>Results of multivariate logistic regression model adjusted for age (continuously), sex, education, physical activity, BMI (categorical), CVD, diabetes. Study participants with other (e.g. VD) or unknown dementia forms were excluded.

**Supplemental Table 15.** Exploratory subgroup and sensitivity analyses for the association of LAP TGF-beta-1 and Alzheimer’s disease

| Group | n<sub>total</sub> | n<sub>cases</sub> | OR (95% CI)<sup>a</sup> | p-value interaction |
| --- | --- | --- | --- | --- |
| **Total cohort** | 1782 | 163 | 1.46 (1.21-1.76) |  |
| **Stratified by age** |  |  |  | 0.6941 |
| <68 years | 1258 | 88 | 1.41 (1.12-1.78) |  |
| ≥68 years | 524 | 75 | 1.49 (1.09-2.03) |  |
| **Stratified by sex** |  |  |  | 0.9685 |
| Women | 965 | 95 | 1.51 (1.17-1.94) |  |
| Men | 817 | 68 | 1.47 (1.09-1.97) |  |
| **Stratified by obesity** |  |  |  | 0.3398 |
| BMI < 30 kg/m<sup>2</sup> | 479 | 53 | 1.30 (0.88-1.93) |  |
| BMI ≥ 30 kg/m<sup>2</sup> | 1303 | 110 | 1.53 (1.23-1.91) |  |
| **Stratified by diabetes** |  |  |  | 0.7049 |
| No | 1489 | 131 | 1.48 (1.20-1.84) |  |
| Yes | 293 | 32 | 1.64 (1.05-2.54)<sup>b</sup> |  |
| **Stratified by CVD** |  |  |  | 0.3792 |
| No | 1373 | 124 | 1.55 (1.25-1.93) |  |
| Yes | 409 | 39 | 1.28 (0.88-1.85)<sup>b</sup> |  |
| **Stratified by APOE ε4** |  |  |  | 0.3987 |
| Negative | 1276 | 87 | 1.59 (1.21-2.07) |  |
| Positive | 506 | 76 | 1.33 (1.02-1.74) |  |
| **Stratified by time of diagnosis** |  |  |  | NA |
| In first 10 years of FUP | 1699 | 80 | 1.47 (1.13-1.91) |  |
| In year 11-19 of FUP | 1702 | 83 | 1.46 (1.15-1.85) |  |
| **Excluding subjects free of dementia who died prior to 80<sup>th</sup> birthday** |  |  |  | NA |
| No | 1782 | 163 | 1.46 (1.21-1.76) |  |
| Yes | 1601 | 163 | 1.56 (1.28-1.90) |  |
| **Excluding subjects with sign of acute infection (CRP level >20mg/L)** |  |  |  | NA |
| No | 1782 | 163 | 1.46 (1.21-1.76) |  |
| Yes | 1745 | 160 | 1.46 (1.21-1.77) |  |

Abbreviations: LAP TGF-beta-1, Latency-associated peptide transforming growth factor beta-1; OR, odds ratio; CI, confidence interval; CVD, cardiovascular disease; APOE, apolipoprotein E; CRP, C-reactive protein;
NOTE: Numbers printed in bold are statistically significant (p < 0.05).
<sup>a</sup>Results of multivariate logistic regression model adjusted for age (continuously), sex, education, physical activity, BMI (categorical), CVD, diabetes, depression, *APOE* genotype. Study participants with other (e.g. VD) or unknown dementia forms were excluded.
<sup>b</sup>Results of multivariate logistic regression model adjusted for age (continuously), sex, education, physical activity, BMI (categorical), CVD, diabetes. Study participants with other (e.g. VD) or unknown dementia forms were excluded.

**Supplemental Table 16.** Exploratory subgroup and sensitivity analyses for the association of VEGF-A and vascular dementia

| Group | n<sub>total</sub> | n<sub>cases</sub> | OR (95% CI)<sup>a</sup> | p-value interaction |
| --- | --- | --- | --- | --- |
| **Total cohort** | 1782 | 195 | 1.43 (1.20-1.70) |  |
| **Stratified by age** |  |  |  | 0.6534 |
| <68 years | 524 | 98 | 1.24 (1.00-1.54) |  |
| ≥68 years | 1258 | 97 | 1.75 (1.31-2.34) |  |
| **Stratified by sex** |  |  |  | 0.7408 |
| Women | 965 | 100 | 1.41 (1.10-1.79) |  |
| Men | 817 | 95 | 1.45 (1.13-1.86) |  |
| **Stratified by obesity** |  |  |  | 0.5126 |
| BMI < 30 kg/m<sup>2</sup> | 479 | 49 | 1.34 (0.95-1.90) |  |
| BMI ≥ 30 kg/m<sup>2</sup> | 1303 | 146 | 1.48 (1.21-1.81) |  |
| **Stratified by diabetes** |  |  |  | 0.7305 |
| No | 1489 | 147 | 1.44 (1.19-1.74) |  |
| Yes | 293 | 48 | 1.40 (0.91-2.16) |  |
| **Stratified by CVD** |  |  |  | 0.1381 |
| No | 1373 | 124 | 1.34 (1.08-1.65) |  |
| Yes | 409 | 71 | 1.74 (1.24-2.44) |  |
| **Stratified by APOE ε4** |  |  |  | 0.5134 |
| Negative | 1276 | 127 | 1.47 (1.19-1.82) |  |
| Positive | 506 | 68 | 1.32 (0.97-1.81) |  |
| **Stratified by time of diagnosis** |  |  |  | NA |
| In first 10 years of FUP | 1666 | 79 | 1.75 (1.33-2.32) |  |
| In year 11-19 of FUP | 1703 | 116 | 1.29 (1.04-1.59) |  |
| **Excluding subjects free of dementia who died prior to 80<sup>th</sup> birthday** |  |  |  | NA |
| No | 1782 | 195 | 1.43 (1.20-1.70) |  |
| Yes | 1601 | 195 | 1.52 (1.27-1.82) |  |
| **Excluding subjects with sign of acute infection (CRP level >20mg/L)** |  |  |  | NA |
| No | 1782 | 195 | 1.43 (1.20-1.70) |  |
| Yes | 1745 | 192 | 1.46 (1.22-1.74) |  |

Abbreviations: VEGF-A, Vascular endothelial growth factor-A; OR, odds ratio; CI, confidence interval; CVD, cardiovascular disease; APOE, apolipoprotein E; CRP, C-reactive protein;
NOTE: Numbers printed in bold are statistically significant (p < 0.05).
<sup>a</sup>Results of multivariate logistic regression model adjusted for age (continuously), sex, education, physical activity, BMI (categorical), CVD, diabetes, depression, *APOE* genotype. Study participants with other (e.g. AD) or unknown dementia forms were excluded.

